# Opening the digital doorway to sexual healthcare: Recommendations from a Behaviour Change Wheel analysis of barriers and facilitators to seeking online sexual health information and support among underserved populations

**DOI:** 10.1101/2024.07.11.24310216

**Authors:** Julie McLeod, Claudia S. Estcourt, Jennifer MacDonald, Jo Gibbs, Melvina Woode Owusu, Fiona Mapp, Nuria Gallego Marquez, Amelia McInnes-Dean, John Saunders, Ann Blandford, Paul Flowers

## Abstract

**Background:** The ability to access and navigate online sexual health information and support is increasingly needed in order to engage with wider sexual healthcare. However, people from underserved populations may struggle to pass though this “digital doorway”. Therefore, using a behavioural science approach, we first aimed to identify barriers and facilitators to: i) seeking online sexual health information and ii) seeking online sexual health support. Subsequently, we aimed to generate theory-informed recommendations to improve these access points.

**Methods:** The PROGRESS framework guided purposive recruitment (October 2021–April 2022) of 35 UK participants from diverse backgrounds, including 51% from the most deprived areas and 26% from minoritised ethnic groups. Semi-structured interviews and thematic analysis identified barriers and facilitators to seeking online sexual health information and support. A Behaviour Change Wheel (BCW) analysis then identified recommendations to better meet the needs of underserved populations.

**Results:** We found diverse barriers and facilitators. Barriers included low awareness of and familiarity with online information and support; perceptions that online information and support were unlikely to meet the needs of underserved populations; overwhelming volume of information sources; lack of personal relevancy; chatbots/automated responses; and response wait times. Facilitators included clarity about credibility and quality; inclusive content; and in-person assistance. Recommendations included: Education and Persuasion e.g., online and offline promotion and endorsement by healthcare professionals and peers; Training and Modelling e.g., accessible training to enhance searching skills and credibility appraisal; and Environmental Restructuring and Enablement e.g., modifications to ensure online information and support are simple and easy to use, including video/audio options for content.

**Conclusions:** Given that access to many sexual health services is now digital, our analyses produced recommendations pivotal to increasing access to wider sexual healthcare among underserved populations. Implementing these recommendations could reduce inequalities associated with accessing and using online sexual health service.

## Introduction

Over the past decade, the online delivery of sexual healthcare has increased, accelerated by the COVID-19 pandemic (1–4). Such healthcare includes online postal self-sampling (OPSS) for sexually transmitted infection (STI) and blood borne virus (BBV) testing (e.g., 5–8). More complex online clinical care pathways are also in development, such as the eSexual Health Clinic for accessing STI test results and treatment (9) and ePrEP for accessing HIV prevention medication, pre-exposure prophylaxis (PrEP) (10). For many people, the initial steps to accessing sexual healthcare, both online and traditional (i.e., in-person/phone), are seeking sexual health information and support online (11–13). Our definition of seeking online sexual health information is inclusive, referring to searching for, finding, understanding, and applying information from the internet (e.g., 14–16) typically found through search engines. Equally, regarding seeking online sexual health support, we refer to finding and using text-based interactions for answers to a range of sexual health queries. These include tools such as live chats and chatbots (synchronous communication with a trained professional (live chats) or with automatic responses (chatbots)) and email or short-messaging service (SMS) text exchange (asynchronous communication with a trained professional (e.g., 17–22). See Appendix A for a list of examples. Together, these two steps (seeking online sexual health information and support) form a digital doorway to wider sexual healthcare (23–27).

Online sexual healthcare can overcome common barriers to accessing traditional sexual health services, offering privacy and convenience (e.g., 12,28,29). However, many may struggle to access and use online sexual healthcare due to inequalities patterned by socio-economic demographics, such as gender, sexual identity, ethnicity, and socio-economic status (3,30–40) (i.e., underserved populations (44)). Further, for people to engage with and pass through the digital doorway to wider sexual healthcare, they require sufficient digital literacy (skills to perform tasks and solve problems in digital environments (41)) and health literacy (capability to understand, evaluate, and use information and services to make choices about health (42)). This complex intersection of socio-economic factors precluding access to healthcare for those who often bear a disproportionate burden of STIs (e.g., 43) illustrates how the provision of online sexual healthcare has the potential to widen inequalities amongst underserved populations.

To prevent widening inequalities in access to online sexual healthcare, it is vital to understand the barriers and facilitators to the digital doorway among underserved populations, theorise the factors that underpin barriers and facilitators, and then identify appropriate theoretically informed recommendations for change (45). While some research exists in this area, the existing literature base is outdated (46–52) or uses exclusively quantitative methods (53–59). Thus, there is an absence of contemporary, in-depth research. Moreover, to our knowledge, there are no studies identifying evidence-based and theoretically informed recommendations for seeking online sexual health information and support among underserved populations. Therefore, using a behavioural science approach, we first aimed to identify barriers and facilitators to two key elements of the digital doorway: i) seeking online sexual health information and ii) seeking online sexual health support. Subsequently, we aimed to propose theory-informed recommendations to improve these two access points. We developed three research questions (RQs):

RQ1) What are the barriers and facilitators to seeking online sexual health information among underserved populations?

RQ2) What are the barriers and facilitators to seeking online sexual health support among underserved populations?

RQ3) What evidence-based and theoretically informed recommendations can be made to enhance seeking online sexual health information and support among underserved populations?

## Methods

### Design

A behaviourally focused cross-sectional qualitative approach, conducted as part of the SEQUENCE Digital Programme (https://www.sequencedigital.org.uk/).

### Applying a behavioural lens

High quality applied behavioural science requires a considered understanding of the specific behaviour(s) that are intended to be changed by an intervention (60). Within the broad behavioural system of ‘accessing and using online sexual healthcare’ we identified seven distinct yet interconnected behavioural domains (see Appendix B). Here, we focus on the first of these two domains that we consider to constitute the digital doorway: 1) seeking online sexual health information and 2) seeking online sexual health support.

### Participants

Using a PROGRESS (Place of Residence, Race/Ethnicity, Occupation, Gender/Sex, Religion, Education, Socio-economic Status, Social Network) (61,62) informed purposive sampling framework (Appendix C), we recruited participants from underserved populations. See Table 1 for participant characteristics and demographics. Representatives from five regional National Health Service (NHS) Trusts/Boards (i.e., organisational areas), two non-governmental organisations (NGOs), and one community college across England and Scotland referred interested potential participants to the research team. One NGO served people with disabilities and learning difficulties, the other, people who identify as LGBTQI+ and Muslim. The community college served people with low educational attainment living in a deprived area. We then telephoned each referred participant to verify eligibility according to our inclusion criteria, screen them against the target sampling frame, collect demographics and information on internet use (see Appendix D), and schedule an interview (phone, video, or face-to-face). Inclusion criteria were: 1) never ordered/used or struggled to order/use an STI self-sampling kit (to recruit participants of lower digital literacy); 2) aged 16+; 3) sexually active; 4) had phone and internet access to enable data collection; 5) lived in the UK; and 6) spoke English well enough to participate in an interview.

**Table 1.** Participant self-reported socio-economic demographic characteristics.

| Variable | n | %<sup>a</sup> |
| --- | --- | --- |
| **Age** |  |  |
| 16-24 | 8 | 22.9 |
| 25-34 | 10 | 28.6 |
| 35-44 | 13 | 37.1 |
| 45-54 | 1 | 2.9 |
| 55-64 | 1 | 2.9 |
| 65+ | 1 | 2.9 |
| **Ethnicity<sup>b</sup>** |  |  |
| Asian, Asian British or Asian Welsh: Chinese | 1 | 2.9 |
| Asian, Asian British or Asian Welsh: Pakistani | 4 | 11.4 |
| Black, Black British, Black Welsh, Caribbean or African: African | 3 | 8.6 |
| Mixed or Multiple ethnic groups: Other Mixed or Multiple ethnic groups | 1 | 2.9 |
| White: Irish | 1 | 2.9 |
| White: English, Welsh, Scottish, Northern Irish, or British | 19 | 54.3 |
| Other self-identified groups (e.g., Hungarian, Italian, Jewish) | 5 | 14.3 |
| **Sexual orientation** |  |  |
| Bisexual (M) | 2 | 5.7 |
| Bisexual (F) | 5 | 14.3 |
| Bi/pansexual/queer (gender-diverse) | 3 | 8.6 |
| Heterosexual/straight (M) | 6 | 17.1 |
| Heterosexual/straight (F) | 10 | 28.6 |
| Homosexual/gay (M) | 7 | 20.0 |
| No response | 1 | 2.9 |
| **Gender** |  |  |
| Cisgender woman | 16 | 45.7 |
| Cisgender man | 16 | 45.7 |
| Gender diverse (non-binary, trans masc) | 2 | 5.7 |
| **Education** |  |  |
| Secondary/High school | 6 | 17.1 |
| College (introductory/foundational vocational award to diploma) | 12 | 34.3 |
| University (Undergraduate student) | 5 | 14.3 |
| University (Undergraduate degree achieved) | 5 | 14.3 |
| University (Postgraduate degree achieved) | 5 | 14.3 |
| No response | 1 | 2.9 |
| **Occupation** |  |  |
| Unemployed | 11 | 31.4 |
| Student (full-time/part-time) | 9 | 25.7 |
| Employed (full-time/part-time) | 13 | 37.1 |
| Retired | 1 | 2.9 |
| **Area of deprivation <sup>c</sup>** |  |  |
| Least deprived (IMD Quintiles 3, 4 and 5) | 14 | 40.0 |
| Most deprived (IMD Quintiles 1 and 2) | 17 | 48.6 |
| No postcode (i.e., homeless) | 1 | 2.9 |
| No response | 2 | 5.7 |
| **Difficulty making ends meet <sup>d</sup>** |  |  |
| Yes | 9 | 25.7 |
| Sometimes | 3 | 8.6 |
| Not anymore but have in the past | 2 | 5.7 |
| No | 20 | 57.1 |
| **Belong to any particular religion or faith** |  |  |
| No (never, not anymore) | 20 | 57.1 |
| Maybe (e.g., spiritual, agnostic) | 2 | 5.7 |
| Yes | 12 | 34.3 |
| *Christianity* | 7 | 20.0 |
| *Islam* | 4 | 11.4 |
| *Judaism* | 1 | 2.9 |
| **Disability** |  |  |
| Learning disability (n=34) | 10 | 28.6 |
| Mental or physical disability (n=33) | 17 | 48.6 |
| *Reduces ability to carry out day-to-day activities* | 12 | 34.3 |
| *Sometimes reduces ability to carry out day-to-day activities* | 1 | 2.9 |
| *Does not reduce ability to carry out day-to-day activities* | 3 | 8.6 |
| **First language** |  |  |
| English | 26 | 74.3 |
| Non-English (Italian, Indonesian, French, Hungarian, Yoruba, Urdu, Gaelic) | 8 | 22.9 |
| **Country born** |  |  |
| UK (England, Scotland) | 26 | 74.3 |
| *England* | 13 | 37.1 |
| *Scotland* | 13 | 37.1 |
| Non-UK (Italy, Indonesia, Switzerland, Hungary, Nigeria) | 8 | 22.9 |

<sup>a</sup>Participant demographics for one participant were not obtained, table includes demographics for n=34, except where participants did not wish to answer the question. Percentages are calculated for N=35.
<sup>b</sup>Reported using the Official National Statistics classifications (68)
<sup>c</sup>Index of Multiple Deprivation (69)
<sup>d</sup>“Making ends meet” = ability to pay for essentials needed to live.

## Materials

We composed a questionnaire-based assessment of eligibility to take part in the study, demographics (based on the PROGRESS framework (61,62)), and experience and skills using technology and the internet (Appendix D). In line with inclusive guidance (63,64), the questions asked regarding socio-economic demographics, such as gender identity and ethnicity, were open questions to capture how participants self-identified.

We also developed an interview topic guide (Appendix E) and supporting visual aids (Appendix F). The topic guide was used to explore participants’ barriers (e.g., what makes it difficult, what are the drawbacks) and facilitators (e.g., what makes it easy, what are the benefits) to both seeking online sexual health information and support. Critically, barriers and facilitators could be either experiential (i.e., actually experienced) or hypothetical. The visual aids depicted the two behavioural domains of seeking online sexual health information and support. Respectively, these visual aids showed search engines with the typed words “sexually transmitted infection (STI) symptoms” for ‘seeking online sexual health information’ and examples of a sexual health live chat and SMS text exchange with a healthcare provider (HCP) for ‘seeking online sexual health support’.

### Participant and patient involvement and engagement

For material development, we consulted public and patient involvement and engagement (PPIE) representatives (N=12) of diverse ages, genders, ethnicities, sexual orientations, religions, and experiences of disability, learning difficulties, and digital STI healthcare. The representatives offered intersectional perspectives and advice on our questionnaire-based assessment of participant demographics and internet use (n=5) and the interview topic guide and visual aids (n=7) to be used within data collection. Consent to share PPIE representatives’ demographic information was not obtained.

### Procedure

One-to-one semi-structured interviews (duration range: 38-82 minutes, *M*=60 minutes) were conducted remotely (video call n=7, phone n=23) or face-to-face (n=5) (all by JMcL). Prior to the interview, we provided participants with an information sheet, consent form, and the visual aids by email or WhatsApp for remote interviews and hard copy for face-to-face interviews. At the beginning of the interview, we obtained verbal consent, recorded via an encrypted recorder for remote interviews and written consent for face-to-face interviews. At the end of the interview, all participants were offered a shopping voucher (value £30) and were provided with a list of sexual health resources (Appendix G) either by email, WhatsApp or hard copy.

### Analysis

For RQs one and two, using NVivo (version 12), we conducted inductive thematic analysis (65) to identify barrier and facilitator themes for each of the behaviours (conducted by JMcL and checked by PF). To do this, we described data using a brief summary barrier or facilitator statement (e.g., “*I suppose, when my son was younger, I worried that he might see…. […] search history and stuff like that, might have put me off*” was described as ‘Concern about family seeing search history’). We then grouped similar summary statements for each behaviour to identify themes (e.g., ‘Concern about family seeing search history’, ‘Having a shared device – lack of privacy’ and ‘Concern about people you know, particularly partners, finding out you may have an STI’ were grouped together and labelled as the theme, ‘Concerns about privacy’).

For RQ three, using the BCW approach (45), barrier and facilitator themes were mapped onto appropriate components of the COM-B Model (66) (by JMcL and checked by PF). The COM-B model posits that behaviour is determined by ‘**Capability**’ (physical and psychological attributes of a person), ‘**Opportunity’** (physical and social attributes of a person’s environment), and/or ‘**Motivation’** (a person’s reflective and automatic mental processes). The COM-B components were then matched to relevant Intervention Functions (nine broad categories of potential interventions to change the capability, opportunity and/or motivation to engage in a behaviour) (45,67) (by PF, audited by JMcL and JMacD). Subsequently, we drew on our collective expertise to operationalise the Intervention Functions into recommendations (conducted by PF and JM, audited by JMacD). The recommendations were reviewed by an interdisciplinary team including sexual health clinicians, public health researchers, and human computer interaction specialists. All BCW analyses were conducted by health psychology researchers who have completed the behaviour change technique taxonomy (version 1) training (https://www.bct-taxonomy.com/). Throughout all stages of analysis, discrepancies were resolved through discussion between JMcL, PF and, latterly, JMacD.

### Ethics

Ethical approval for this study was granted by the East of England - Cambridge South Research Ethics Committee (REC) (reference 21/EE/0148) and Glasgow Caledonian University REC (reference HLS/NCH/20/045).

## Results

### Participants

Participants (N=35) (see Table 1) ranged in age from 18-70 (*M*=34 years) and were diverse, representing several underserved populations: 51% (n=18) lived in the most deprived areas of the UK; 51% (n=18) had no higher (i.e., university) education; 40% (n=14) were of a minoritised ethnic group, of which, five were from an ethnic group other than White; 23% (n=8) did not speak English as their first language; 49% (n=17) had a mental or physical illness or condition lasting 12 months or more; and 29% (n=10) had a learning difficulty.

The majority of participants reported owning a digital device (e.g., mobile phone or laptop) to access the internet (n=30, 86%) and using the internet every day (n=29, 83%) for a wide range of activities, most frequently, social media (n=22, 63%), work including research (n=17, 48%), streaming TV shows or videos (n=10, 29%), searching the internet for information (n=9, 26%), and checking news (n=5, 14%) and emails (n=5, 14%). Over half of the participants described themselves as having ‘high’ level skills using the internet (i.e., digital literacy) (n=20, 57%), 12 reported ‘medium’ (34%) and 2 reported ‘low’ (6%). Over a third (n=12, 34.3%) had never searched for any sexual health information online, none had ever used a live chat or email or text exchange service for getting sexual health support, and few had used other online sexual health services such as booking an appointment (n=4, 11.4%) or ordering medication through a private (non-state-funded) clinic (n=1, 2.9%). Moreover, the majority had never ordered a postal STI self-sampling kit (n=24, 68.6%) and a few reported having tried and struggled to order (n=3, 9%), or use (n=7, 20%), a self-sampling kit for STIs and blood borne viruses. See Appendix H for an overview of participant data regarding experience and self-rated skills of using the internet.

### RQ1) What are the barriers and facilitators to seeking sexual health information online among underserved populations?

Table 2 details the nine barrier and eleven facilitator themes to seeking online sexual health information, with indicative data extracts, and their corresponding COM-B components.

**Table 2.**
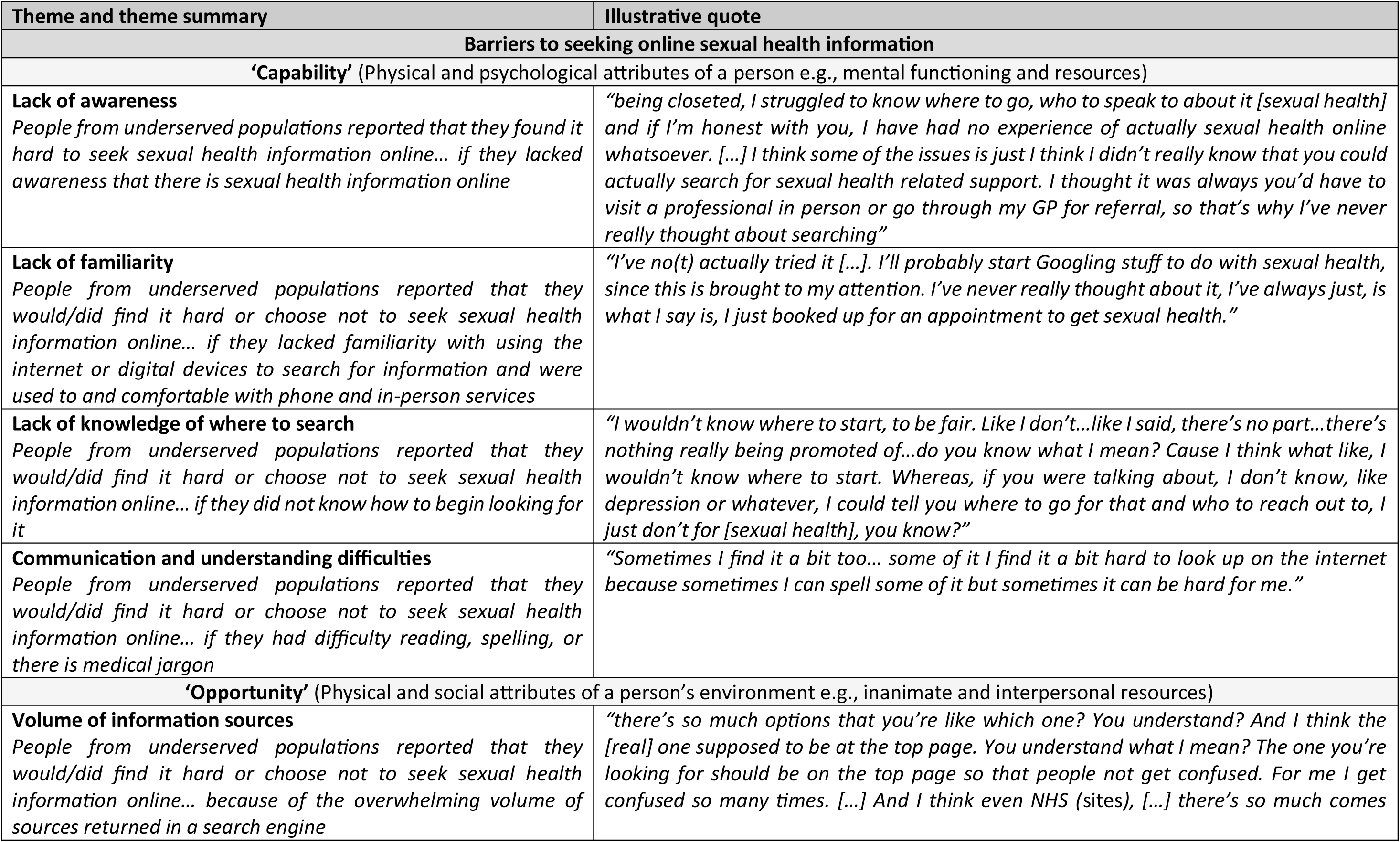
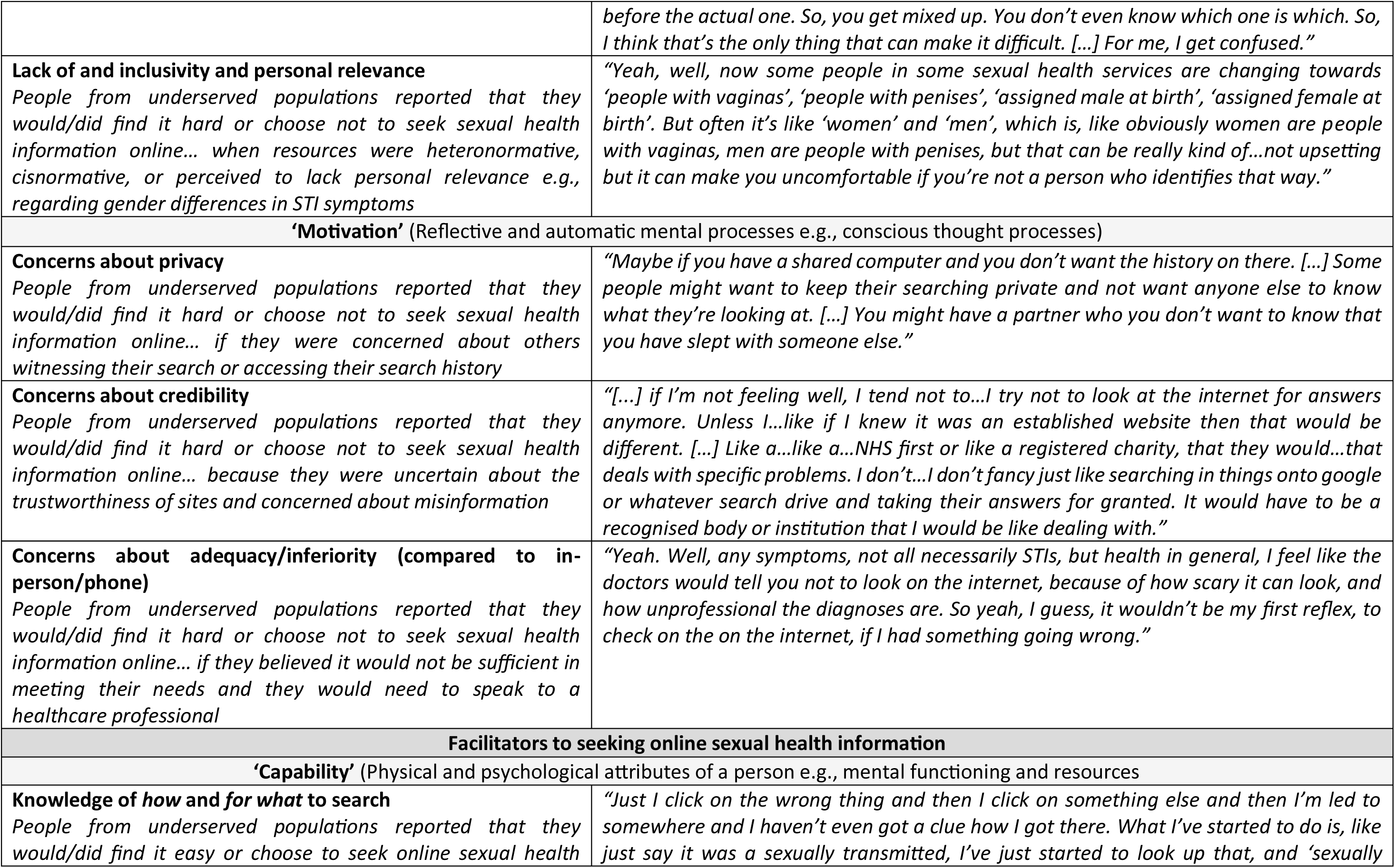
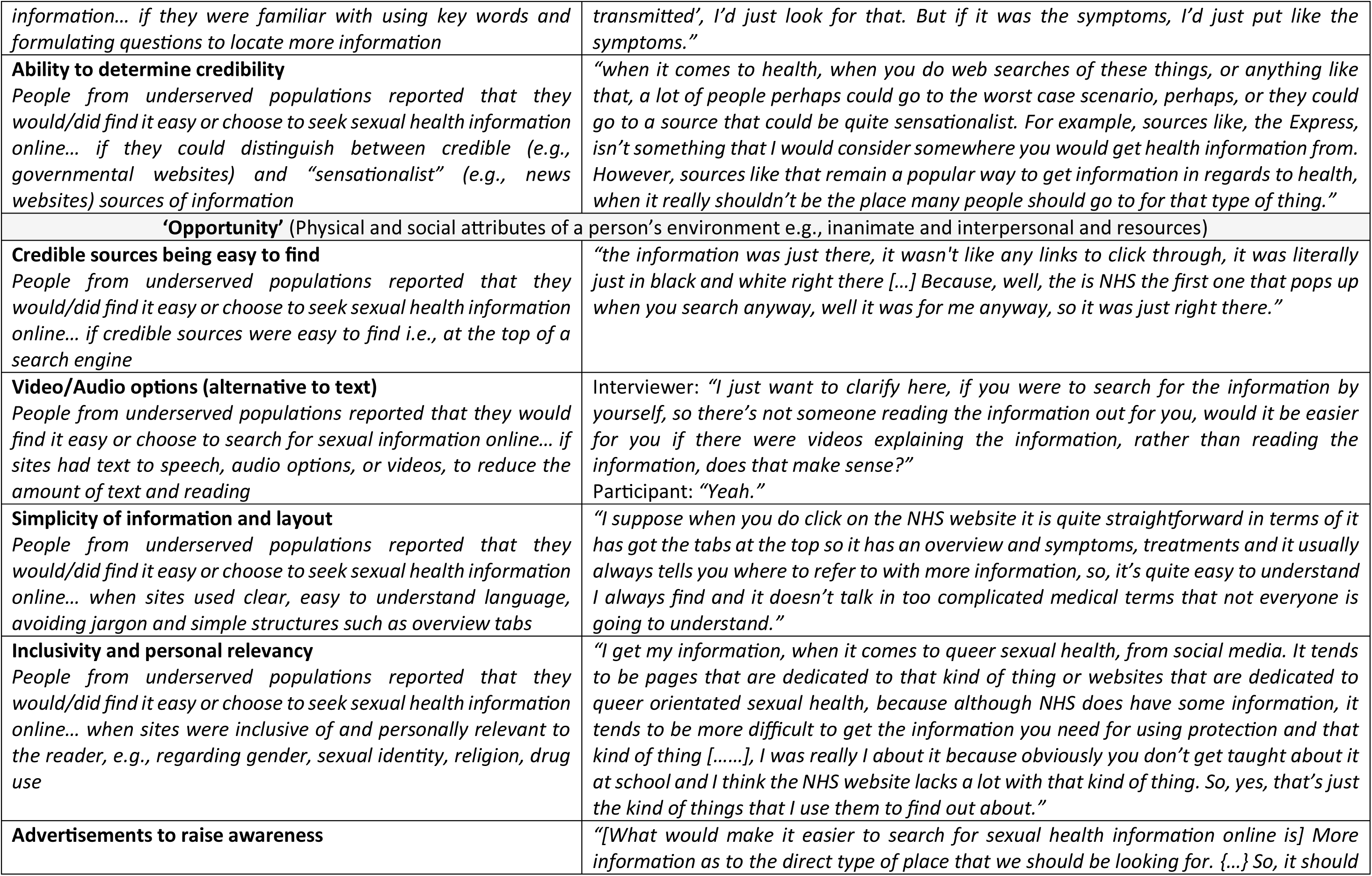
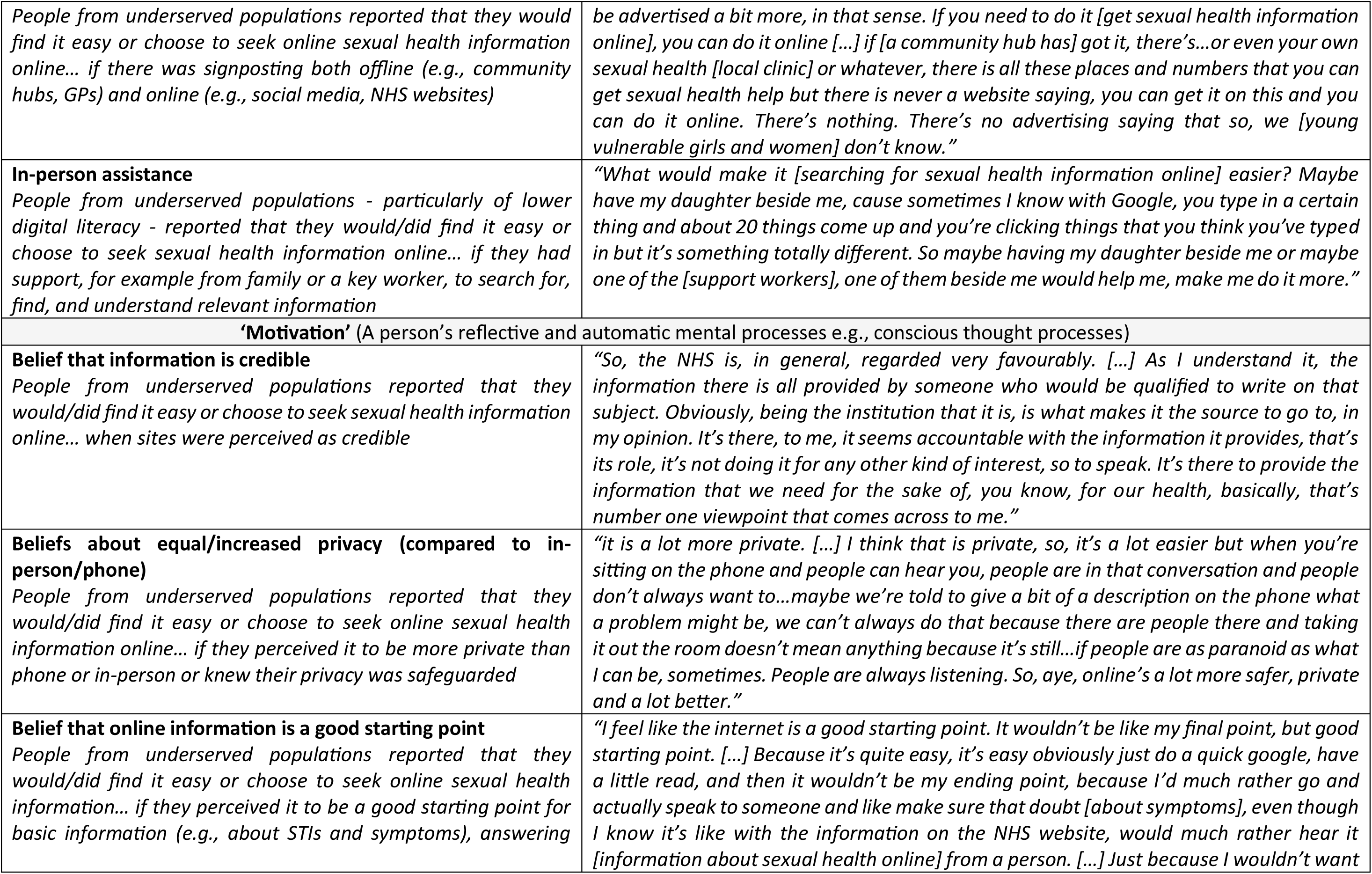
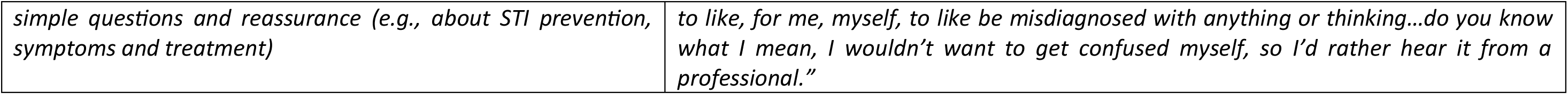
Barrier and facilitator themes and related COM-B components for seeking online sexual health information among underserved populations.

Barrier themes relating to **‘Capability’** were ‘*Lack of awareness’; ‘Lack of familiarity’; ‘Lack of knowledge of where to search’;* and ‘*Communication and understanding difficulties’.* This cluster of barriers related to a lack of important knowledge about the existence of sexual health information and how to access and understand it. The barrier theme relating to **‘Opportunity’** focussed on features of the digital environment: ‘*Volume of information sources’* and *‘Lack of inclusivity and personal relevance’.* This barrier provides a sense of how some people find seeking online sexual health information overwhelming whilst navigating search engine results and finding relevant information. Barrier themes relating to **‘Motivation’** were negative perceptions of online sexual health information: ‘*Concerns about privacy’; ‘Concerns about credibility’; ‘Concerns about adequacy/inferiority (compared to in-person/phone)’.* This cluster related to concerns about the internet as a source of sexual health information.

Facilitator themes related to **‘Capability’** centred around knowledge: ‘*Knowledge of how and for what to search’* and ‘*Ability to determine credibility’.* Facilitator themes related to **‘Opportunity’** focussed on features of the digital environment, promotion, and help: ‘*Credible information being easy to find’; ‘Video and audio options (alternative to text)’; ‘Simplicity of information and layout’; ‘Inclusivity and personal relevancy’; ‘Advertisements to raise awareness’;* and *‘In-person assistance’.* Further, in-person assistance speaks to the need for help to seek online sexual health information, for example from a key worker or family member, among those of lower digital literacy or with learning difficulties. Facilitator themes relating to **‘Motivation’** were positive perceptions on online sexual health information: ‘*Belief that information is credible’; ‘Beliefs about equal/increased privacy (compared to in-person/phone)’;* and ‘*Belief that online information is a good starting point’*.

### RQ2) What are the barriers and facilitators to seeking online sexual health support among underserved populations*?*

Table 3 details eight barrier and eight facilitator themes to seeking online sexual health support (via a synchronous live chat or asynchronous email or SMS text exchange), with illustrative extracts, and corresponding COM-B components.

**Table 3.** Barrier and facilitator themes and corresponding COM-B components for seeking online sexual health support among underserved populations.

| Theme and theme summary | Indicative quotes |
| --- | --- |
| **Barriers to seeking online sexual health support** |  |
| **‘Capability’** (Physical and psychological attributes of a person e.g., mental functioning and resources) |  |
| **Lack of awareness**<br>*People from underserved populations reported that they would find it hard to get sexual health support online... because they lacked awareness that online sexual health support exists* | *“If I’m being honest, I didn’t even know they existed, the live chats, through the sexual health worker. [...] I’m one of the older generation, though I’m 61, so I’m kind of older and I actually just even got a smartphone and I’m not clued into all that. I’ve come across a few people that have not got any computer skills at all. At least I’ve got some.”* |
| **Lack of familiarity**<br>*People from underserved populations reported that they would find it hard or choose not to get sexual health support online... if they lacked familiarity with using a live chat, email, or text exchange service and were used to phone or in-person services* | *“I’ve used services through calling the services, and that’s what I’ve always known. Because I’ve not used live chat services like that, I wouldn’t know, so I wouldn’t know what would be involved”* |
| **‘Motivation’** (A person’s reflective and automatic mental processes e.g., conscious thought processes) |  |
| **Concerns about confidentiality and anonymity**<br>*People from underserved populations reported that they would find it hard or choose not to get sexual health support online... if they were concerned about the confidentiality of live chats and worried about others, such as partners or parents, finding out they may have STI or sexual health questions or concerns* | *“I mean as much as I think it’s a good thing, I do...I would still have some concerns about putting in my full...you know, like my real name and stuff like that unless I really knew for sure I was dealing with a confidential company...or like the NHS”* |
| **Concerns about the impersonal nature of online support**<br>*People from underserved populations reported that they would find it hard or choose not to get sexual health support online... if they perceived it to be impersonal and lacking capacity to provide reassurance* | *“Mhm. I guess, my first thought would be, how impersonal it can be. [...] I would say, it’s more of a disadvantage, as in, yeah, it does feel a bit impersonal, and I feel, because of how personal sexual health is, that would be a bit...like a gap...that it’s weird for me.”* |
| **Concerns about understanding and communication**<br>*People from underserved populations reported that they would find it hard or choose not to get sexual health support online... if they had difficulty with spelling or reading and/or were concerned about staff not being able to communicate clearly and effectively* | *“I’d probably be concerned that...whether I was able to understand what was being said in the first place.”* |
| **Concerns about online support meeting their needs**<br>*People from underserved populations reported that they would find it hard or choose not to get sexual health support online... if they believed it would not be sufficient in meeting their needs and they would need to see a healthcare professional in-person* | *"Yeah, yeah, yeah, so as in like, if I was texting someone and I thought I had an STI, it would for me it would just be the same as Googling. I'd personally want to speak to someone physically to kind of possibly be reassured. You can't translate that on a message, you know?"* |
| **Concerns about chatbots and automated responses**<br>*People from underserved populations reported that they would find it hard or choose not to get sexual health support online... if they knew or thought it was being delivered via automated responses instead of a qualified professional or trained member of staff due to concerns about the quality of the responses* | *"I don't really mind if it's trained staff or a doctor or a nurse, but I think I'd... not do it if it's a chatbot, because sometimes they can be not helpful. [...] Because... well, the chatbot I had experienced [for non-sexual-health-related help] only had like set responses and sometimes when you want to use the live chat you kind of have more complicated stuff to ask about, sometimes the set responses aren't that helpful, so it's more frustrating than just not doing it. [...] like I could just Google it instead of talking to a chatbot, cause like I said, if I'm going to do a live chat, it's probably something more complicated and it's like something I can't Google. [...] I'd use like a live chat or an email thing, if it's a healthcare professional on the other end."* |
| **Concerns about response wait times**<br>*People from underserved populations reported that they would find it hard or choose not to get sexual health support online... if there was a wait time for receiving response including hours to days via asynchronous communication or half an hour for synchronous communication* | *"I just probably wouldn't use that because it's just prolonging like whatever problem I need help with. [...] if I wanted to speak to someone and wanted to get help, then it doesn't really seem useful to be waiting for them a couple of days or a day or so for an answer, then send one back and then wait another day, it just doesn't really seem like...don't really see the point in it."* |
| **Facilitators to seeking online sexual health support** |  |
| **'Capability'** (Physical and psychological attributes of a person e.g., mental functioning and resources) |  |
| **Familiarity with online text-based support**<br>*People from underserved populations reported that they would find it easy or choose to get sexual health support online... if they had familiarity with using live chats, email, or text exchange services as modes of communication* | *"If you're very familiar with texting, and using your email, to me, it would be pretty convenient to use it, because obviously you're very familiar with that already."* |
| **'Opportunity'** (Physical and social attributes of a person's environment e.g., inanimate and interpersonal and resources) |  |
| **Immediate responses**<br>*People from underserved populations reported that they would find it easy or choose to get sexual health support online if responses were quick (i.e., immediate), as sexual health queries were typically perceived to be urgent* | *"I imagine it [sexual health live chat] would be quite good cos it would be like immediate...you're immediately directed to someone, so you can ask questions."* |
| **Clear trustworthiness and high quality**<br>*People from underserved populations reported that they would find it easy or choose to get sexual health support online... if they knew the interaction was with a qualified/trained person and, for some, if it was a safe space for discussing LGBTQI+ issues* | *"If I knew this person was working for the NHS, I would... I immediately have a trust. I just trust them completely. I know sometimes there are breaches in confidentiality in all organisations, but I just immediately feel I can trust this person with very sensitive information. If it's a private company, I wouldn't feel the same [...] I wouldn't automatically trust them"* |
| **Personal feel**<br>*People from underserved populations reported that they would find it easy or choose to get sexual health support online... if it felt personal (e.g., had a friendly face, welcome invitation, and/or feels like a chat rather than formal)* | *"someone who kind of talks in their dialect so every time you come on to these chats it's a different speech pattern, it's a different stress pattern on words and you can see it there visually in front of you so you know it's someone different and you know it's a person, if that makes sense. [...] So, just having like a true voice, like allowing people to have that true voice, let the informality, let the slang be there... Maybe give someone, maybe like at the start when opting in for the chat, give someone the option like would you like a more formal chat or an informal chat or if there's another way to discuss that or ask that question."* |
| **Advertisements to increase awareness**<br>*People from underserved populations reported that they would find it easy or choose to get sexual health support online... if there was promotion both offline (e.g., community hubs, GPs) and online (e.g., social media, NHS websites)* | *"Again, like I said, advertisement, more people talking about it [sexual health live chat services], like sexual health clinics and people opening up and talking about it but it's no...if we go to a sexual health team, we never hear anybody, you know you can go online and get this, we don't hear anything like that. It's just, right, see you next week"* |
| **In-person assistance**<br>*People from underserved populations reported that they would find it easy or choose to get sexual health support online... if they had help, e.g., from key workers, to find online services, type questions, and interpret responses* | *"Some people with disabilities who need...who get support would need to do it with support 'cause like me, probably wouldn't understand the long jargon words. And need help to spell"* |
| **'Motivation'** (A person's reflective and automatic mental processes e.g., conscious thought processes) |  |
| **Beliefs about equal/increased privacy (compared to in-person/ phone)**<br>*People from underserved populations reported that they would find it easy or choose to get sexual health support online... if they perceived it to be more private than phone or in-person or knew their privacy was safeguarded* | *"it can feel a bit uncomfortable or a bit embarrassing going to a sexual health clinic in a way. So, it [a sexual health live chat/email/text service] would take that out of it as well. [...] I suppose it's because sexual health is still not something we just talk about openly, especially not as an Asian person. In the Asian community, we don't really talk about sex a lot, so I feel quite like I'm being watched when I do go into the clinic."* |
| **Beliefs about online support meeting their needs**<br>*People from underserved populations reported that they would find it easy or choose to get sexual health support online... if* | *"as far as I understand, these live chat services [...] is to kind of get sexual health support and advice so, and what I understand by sexual health support could be, again, general information about kind of sexual health in general but also kind of support about, say,* |
| *they perceived it to be sufficient to meet their needs, i.e., for general or specific queries, link to in-person services, to arrange a call back, help to order an OPSS, or to book an appointment* | *suppose I might just, I've just had sex and I'm concerned about this...information about a specific instance, for example, am I at risk, am I a risk? So, again, I don't think speaking to someone in person, for example, wouldn't necessarily be better than using the live chat. [...] so if it's just concerning a) some general questions about, say, what kind of services are available or b) general information about STIs and sexual health in general and c) information about...again, suppose I just had a sexual encounter that was deemed to be kind of a risk and I wanted to kind of speak to someone about it because I had some concerns then an online live chat would do absolutely fine."* |

Barrier themes relating to **‘Capability’** were*: ‘Lack of awareness’* and ***‘****Lack of familiarity’* with online text-based support. These themes revealed a lack of awareness of the existence of online sexual health support services and experience using digital technology and the internet for sexual health support. Barrier themes relating to **‘Motivation’** were negative perceptions about digital sexual health support: ‘*Concerns about confidentiality and anonymity’; ‘Concerns about the impersonal nature of online support’; ‘Concerns about understanding and communication’; ‘Concerns about online support meeting their needs’; ‘Concerns about chatbots and automated responses’;* and *‘Concerns about response wait times’.* This cluster of themes highlighted the span of participants’ concerns about seeking online support. Participants consistently reported that they would not use online support for sexual health that had any wait times (i.e., was not instantaneous) to receive a response, or if they would be speaking to a ‘chatbot’ with pre-set or automated responses instead of a person, particularly a trained professional. No barriers corresponded to **Opportunity**.

One facilitator theme related to **‘Capability’** was identified regarding knowledge: ***‘****Familiarity with online text-based support services’.* Facilitator themes corresponding to **‘Opportunity’** related to advertisements, help, and features of the digital environment: ***‘****Immediate responses’; ‘Clear quality and trustworthiness’; ‘Personal feel’; ‘Advertisements to increase awareness’;* and’ *In-person assistance’.* Immediacy of the response was a feature that people felt they needed to engage with online support. Advertisements were also important in relation to ways in which awareness and skills with online support could be enhanced. Further, ‘*in-person assistance’* illustrates the needs of participants of lower digital literacy and/or those with a learning difficulty for help engaging and accessing online support. Facilitator themes relating to **‘Motivation’** were positive perceptions on online sexual health support: ‘*Beliefs about equal/increased privacy (compared to in-person/phone)’* and *‘Beliefs about online support meeting their needs’*.

### RQ3) What evidence-based and theoretically informed recommendations can be made to enhance seeking online sexual health information and support among underserved populations?

The BCW analysis identified potentially useful Intervention Functions to assist underserved populations in seeking online sexual health information and support: **Education, Persuasion, Training, Modelling, Enablement,** and **Environmental Restructuring** (see Appendix I for definitions). From these Intervention Functions, we proposed a range of more specific recommendations outlined below. Table 4 and Table 5 provide an overview of the BCW analysis of barriers and facilitators and proposed recommendations for online sexual health information and support, respectively.

**Table 4.**
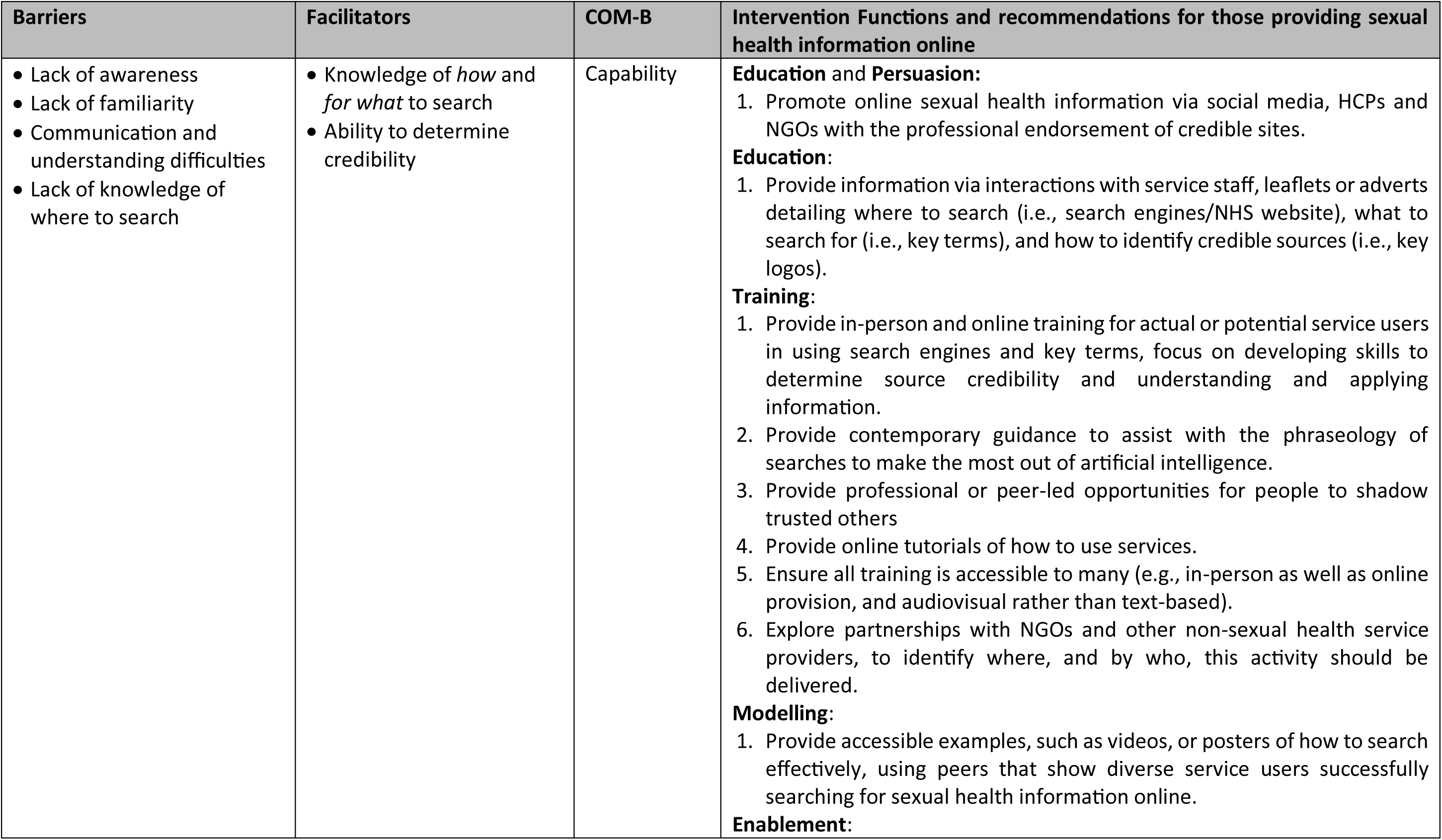
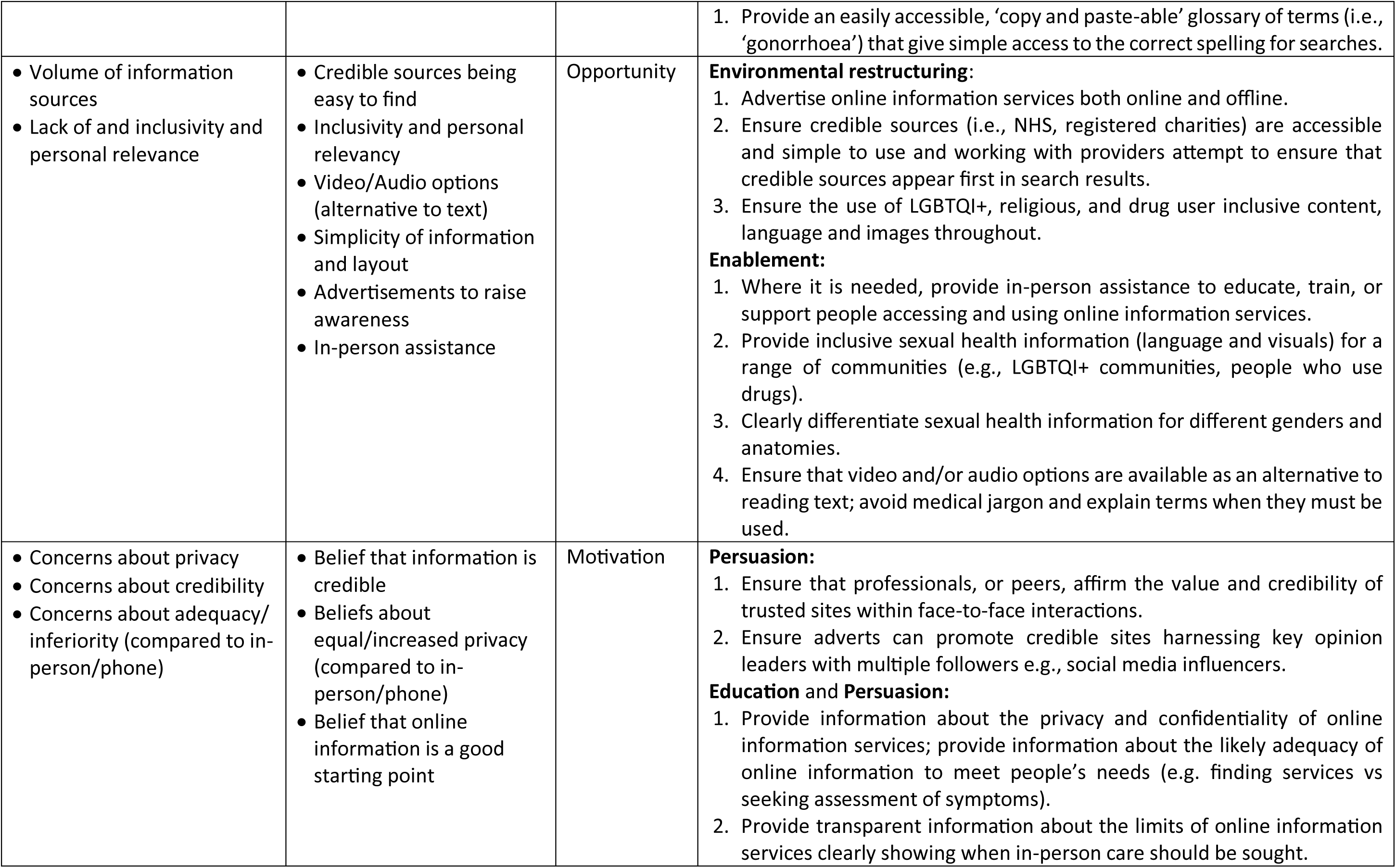
Intervention content from Behaviour Change Wheel to address barriers and facilitators to seeking sexual health information online.

**Table 5.**
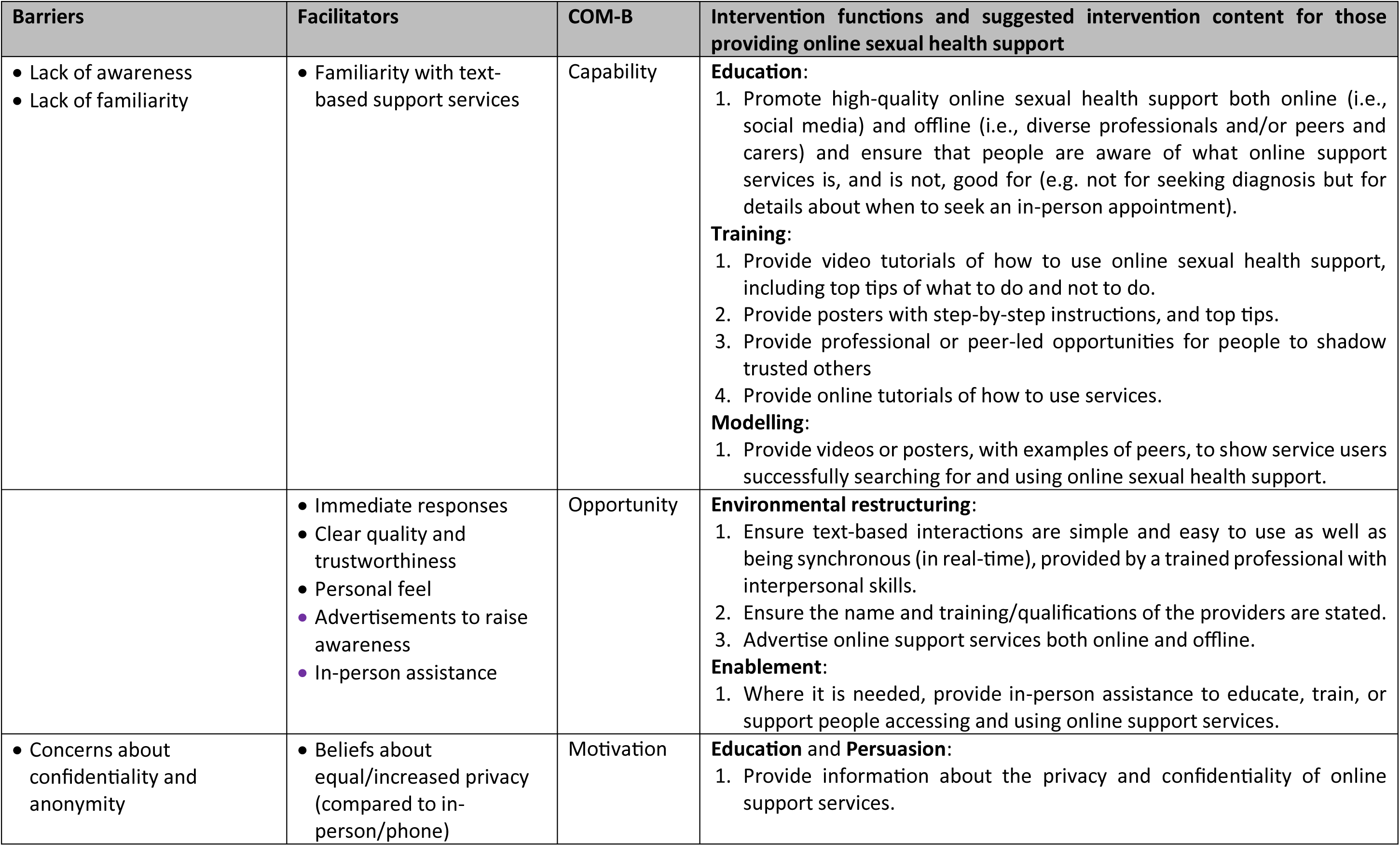
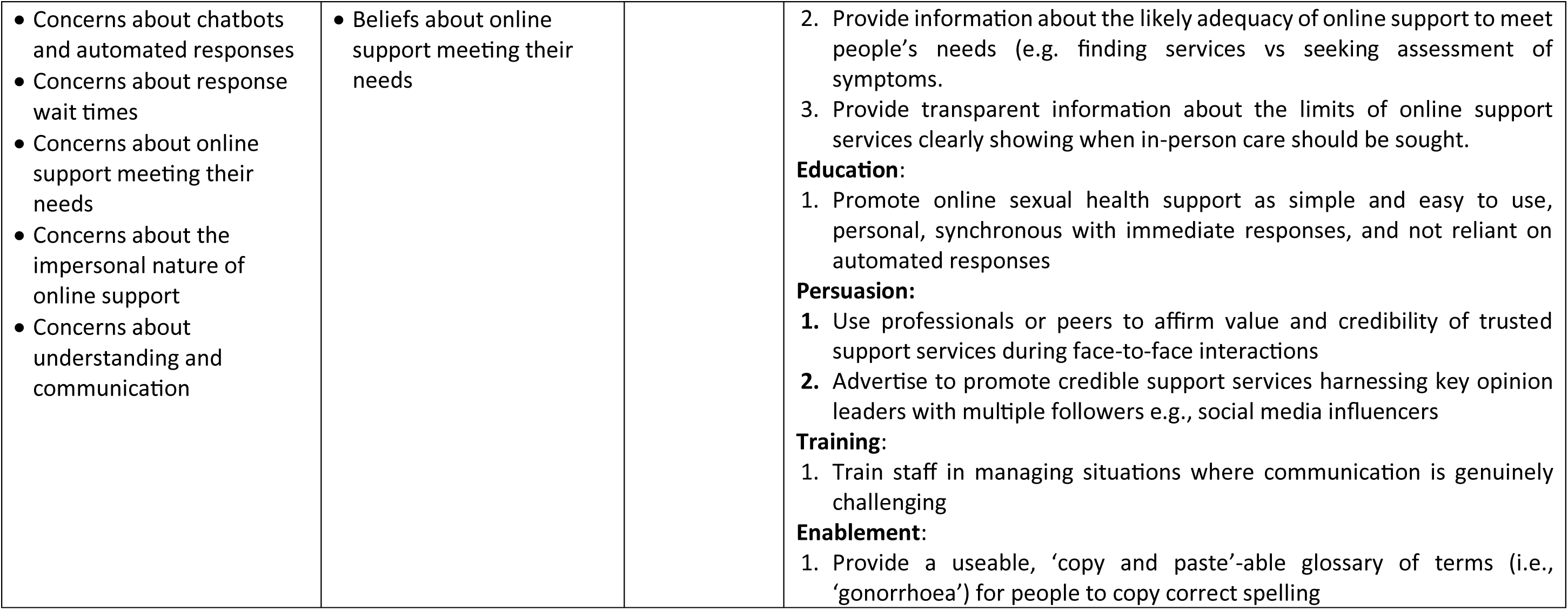
Intervention content from Behaviour Change Wheel to address barriers and facilitators to seeking online sexual health support.

| Barriers | Facilitators | COM-B | Intervention functions and suggested intervention content for those providing online sexual health support |
| --- | --- | --- | --- |
| • Lack of awareness<br>• Lack of familiarity | • Familiarity with text-based support services | Capability | **Education:**<br>1. Promote high-quality online sexual health support both online (i.e., social media) and offline (i.e., diverse professionals and/or peers and carers) and ensure that people are aware of what online support services is, and is not, good for (e.g. not for seeking diagnosis but for details about when to seek an in-person appointment).<br>**Training:**<br>1. Provide video tutorials of how to use online sexual health support, including top tips of what to do and not to do.<br>2. Provide posters with step-by-step instructions, and top tips.<br>3. Provide professional or peer-led opportunities for people to shadow trusted others<br>4. Provide online tutorials of how to use services.<br>**Modelling:**<br>1. Provide videos or posters, with examples of peers, to show service users successfully searching for and using online sexual health support. |
|  | • Immediate responses<br>• Clear quality and trustworthiness<br>• Personal feel<br>• Advertisements to raise awareness<br>• In-person assistance | Opportunity | **Environmental restructuring:**<br>1. Ensure text-based interactions are simple and easy to use as well as being synchronous (in real-time), provided by a trained professional with interpersonal skills.<br>2. Ensure the name and training/qualifications of the providers are stated.<br>3. Advertise online support services both online and offline.<br>**Enablement:**<br>1. Where it is needed, provide in-person assistance to educate, train, or support people accessing and using online support services. |
| • Concerns about confidentiality and anonymity | • Beliefs about equal/increased privacy (compared to in-person/phone) | Motivation | **Education and Persuasion:**<br>1. Provide information about the privacy and confidentiality of online support services. |
| • Concerns about chatbots and automated responses<br>• Concerns about response wait times<br>• Concerns about online support meeting their needs<br>• Concerns about the impersonal nature of online support<br>• Concerns about understanding and communication | • Beliefs about online support meeting their needs |  | 2. Provide information about the likely adequacy of online support to meet people's needs (e.g. finding services vs seeking assessment of symptoms).<br>3. Provide transparent information about the limits of online support services clearly showing when in-person care should be sought.<br>**Education:**<br>1. Promote online sexual health support as simple and easy to use, personal, synchronous with immediate responses, and not reliant on automated responses<br>**Persuasion:**<br>1. Use professionals or peers to affirm value and credibility of trusted support services during face-to-face interactions<br>2. Advertise to promote credible support services harnessing key opinion leaders with multiple followers e.g., social media influencers<br>**Training:**<br>1. Train staff in managing situations where communication is genuinely challenging<br>**Enablement:**<br>1. Provide a useable, 'copy and paste'-able glossary of terms (i.e., 'gonorrhoea') for people to copy correct spelling |

### Education and Persuasion

It may be critical to educate people from underserved populations that online sexual health information and support services exist, for example, through advertising online (e.g., via social media) and offline (e.g., via HCPs and NGOs), and persuade them to use these via endorsement by a range of credible professionals and peers. This education could be delivered via face-to-face interactions with service staff (e.g., HCPs and key workers), posters or leaflets, or posts on professional social media accounts (e.g., from HCPs or influencers) detailing where to search (i.e., search engines/NHS website), what to search for (i.e., key terms), and how to identify credible sources (i.e., key logos). It may also be important to inform people about the benefits and limits of online sexual health information and support and when in-person care should be sought i.e., that online information and support are appropriate for simple issues, such as symptom checking, finding clinics, booking appointments, and signposting, and not appropriate for complex healthcare, such as diagnosis. Further, education regarding the privacy and confidentiality of online sexual health information and support may be important, as well as promoting them as simple and easy to use, personal, synchronous with immediate responses, and not reliant on automated responses). It is critical that Education and Persuasion are delivered alongside Environmental Restructuring and Enablement to ensure online sexual health information and support are private/confidential and simple to use.

### Training and Modelling

Training opportunities for necessary skill acquisition could be part of all services that offer online information and support. Specifically, this could involve step-by-step instructions on using search engines, identifying and typing key terms, determining source credibility, and understanding and applying information. Training could be delivered in the form of videos, posters or leaflets, or professional or peer-led tutorials. To ensure accessibility in relation to digital provision, audiovisual training options should be offered alongside text-based and training should be provided both in-person as well as online. It might also be critical to provide modelling of peers successfully using online sexual health information and support, such as showing videos or images of peers from underserved populations searching for sexual health information such as STI symptoms or using a sexual health live chat to discuss safe sex. Further, online sexual health information and support service providers could explore partnerships with NGOs for delivery of training.

### Environmental restructuring and Enablement

Restructuring the online environment and enabling people from underserved populations to seek online sexual health information and support is essential. Particularly, it may be vital to ensure that online sexual health information and support services are clearly labelled as delivered by credible, trusted sources (i.e., NHS, registered charities) and are designed to be simple to use, avoiding medical jargon and explaining terms when they must be used. Further, enablement could involve providing a glossary of sexual health terms for examples of correct spelling and video and/or audio options for information. Online sexual information and support content could also be demonstrably tailored to a range of communities (e.g., gender, sexuality, and religious minorities, and people who use recreational drugs). Further, in-person assistance could enable people of lower digital literacy and with learning difficulties to access and use sexual health information and support. Finally, online sexual health information and support service providers could ensure that any text-based interactions are synchronous (in real-time) and delivered by a trained professional, stating the name and training/qualifications of the professional involved.

## Discussion

This study is the first to detail barriers and facilitators to seeking online sexual health information and support amongst underserved populations and propose specific recommendations to enable underserved populations to find and use online sexual health information and support. These findings can be applied existing and novel online sexual health information and support services to improve access to this digital doorway to wider sexual health services.

### Barriers and facilitators to passing through the digital doorway

Key barriers to people from underserved populations seeking online sexual health information and support were a lack of awareness of their availability and familiarity with using them; privacy concerns; and the perception of as inadequate to meet varied and complex sexual health needs and inferior to traditional information and support services (e.g., in-person appointment or phone call with a HCP). For seeking online sexual health information, specifically, barriers were navigating the overwhelming volume of information and different sources of and the perceived lack of inclusivity and relevance of information, particularly on government websites. For seeking online sexual health support, specifically, barriers were chatbots and automated responses; asynchronous communication; and wait times for responses. An important consideration here is that none of the participants had ever used an online sexual health support service, despite their availability (e.g., 17–22), thus these barriers were exclusively hypothetical.

Important facilitators to seeking online sexual health information and support were the perceived benefit of increased privacy compared to traditional services; the provision of video and/or audio options as alternatives to text; the presentation of information and support in a simple way – such as step by step and without jargon; and in-person assistance, for example, from key workers, family members, NGO staff, or GPs. For seeking online sexual health information, specifically, facilitators were inclusivity and personal relevancy of sexual health information, particularly for people from LGBTQI+ populations, religious backgrounds, and recreational drug users. For seeking online sexual health support, specifically, facilitators were synchronous communication; the perception of online support as acceptable for simple tasks such as appointment booking, general sexual health advice, and signposting to services; and delivery by qualified personnel.

Some of these findings are novel to this study. Importantly, low awareness of online sexual health information and support was common, even among participants who reported using the internet ‘all the time’. Additionally, the perceived inferiority and inadequacy of online sexual health information compared to traditional sources of sexual health information and unanimous rejection of chatbots and automated responses for sexual health support, due to the perceived complexity of sexual health, were critical. However, beliefs about the inferiority of online services may change, as generative artificial intelligence becomes more commonplace and trusted (80). Further, many of our barrier and facilitators findings are consistent with previous literature, offering a novel perspective from a diverse sample of underserved populations. Privacy benefits and concerns (38,57,71–74) and the value of video and audio options and simplicity (e.g., 29,38,75) have been previously reported amongst a range of populations. Further, the overwhelming volume of sources and information, need for inclusivity and personal relevancy of information, and preferences for online support to be delivered synchronously have been reported previously among sexual minority women (76) and African American youth (75), and young people (38,77,78). Moreover, concerns about the adequacy of online information and support has been reported previously (73,74,79) and the acceptability of online support for simple tasks is in line with research regarding HCPs’ views of chatbots for sexual health (73). Overall, the consistency of these findings with research with other populations indicates that addressing the barriers and enhancing the facilitators from this current study will improve online sexual health information and support for many beyond this sample.

### Recommendations for the digital doorway to wider sexual healthcare

We identified a range of theory informed Intervention Functions to improve access to the digital doorway to wider sexual healthcare, including **Education, Persuasion, Training, Modelling, Environmental Restructuring,** and **Enablement.** First, advertising and promotion of services that provide online sexual health information and support and HCP’s endorsing online services and supporting patients to use them is vital. Together, these findings resonate with wider research on telehealth *per se* (81), highlighting the importance of marketing and communication activities to increase awareness of online services to enable equitable access. Additionally, interventions could include online services offering audio-visual forms of communication, such as text-to-speech or cartoon animations. Moreover, our recommendations include the provision of inclusive and personally relevant information and support, such as information about sexual health relevant to those of diverse sexual and/or gender identities, religious backgrounds, and drug use. This is in line with previous research showing that tailoring of information can positively influence acceptability of interventions and address barriers to care (82–84). Further, interventions could include training HCP’s on how to use and promote online sexual health services effectively, in line with previous research regarding reducing digital inequalities (81). The latter is essential for effective professional endorsement of high-quality online services for sexual health information and support (85). Such endorsements should stress the services’ credibility, useability, and details of its functionality (e.g., trained professional will interact in real time). Interventions could also include embedding optional training for service users regarding how to use online services.

While the BCW analysis identified the above recommendations, it is important to consider that intervention development guidance highlights the centrality of the involvement of relevant stakeholders to co-produce culturally appropriate intervention content (86), whilst considering affordability, practicability, efficacy, acceptability, side-effects, and equity (45). Such stakeholder engagement must consider and address the operationalisation of our proposed recommendations to ensure final intervention content aligns with local circumstances and wider legislative socio-cultural contexts. Furthermore, the involvement of stakeholders in shaping future interventions can help secure their effective delivery, given their investment of time and resource.

### Implications

Changes to the ways online sexual health information and support are provided are required to facilitate more people to pass through the digital doorway to wider sexual healthcare. Given that some of these findings are already well documented in the literature, it is concerning that sufficient changes have not yet been made. Our evidence-based and theoretically informed recommendations (i.e., Tables 5 and 6) should assist these changes to take place. As such, our detailed specification of recommendations is a valuable contribution to the field of sexual health with the potential to catalyse change in the delivery of sexual health information and support online.

### Strengths and limitations

First, the PPIE activities with diverse representatives ensured the development of inclusive study materials that were fit for purpose. Second, as the study was conducted in the UK, some of the findings may be specific to the UK health setting. However, most of the findings presented here are likely to be generalisable to the wider population and to other countries and settings. Third, in addition to a rigorous thematic analysis, we conducted a BCW analysis to provide theory informed and evidence-based recommendations for improvements to seeking online sexual health information and support. As the BCW draws on the cumulative learning from multiple theories of behaviour change (66), it enabled us to specify potentially useful intervention content beyond that suggested by our participants.

Furthermore, our sample included people from a range of underserved populations whose needs are typically unmet by existing services and whose perspectives are often overlooked, such as people from minoritised sexual, ethnic, and religious groups, and those living in the most deprived areas. However, despite our best efforts, we were unable to meet our target recruitment for participants who are from minoritised gender and ethnic groups, particularly those of Black African and Black Caribbean ethnicity who experience a disproportionate burden of STIs (35,43) and have been historically underserved. Such populations have been involved in previous research into online sexual health information and support (48,75,87–89), however, all of these studies were conducted in the United States. Future research should prioritise recruiting people from minoritised gender and ethnic groups in the UK.

## Conclusion

This study identified key barriers and facilitators to seeking online sexual health information and support among a diverse sample of underserved populations. Our BCW analyses then suggested an array of potentially useful changes that could be made to reduce barriers and enhance facilitators to passing through this digital doorway and subsequently increase access to wider in-person and online sexual healthcare. Overall, our recommendations focus on adding to existing services in ways that enable used of them and educate, persuade, and offer accessible training to those from underserved populations. Recommendations include educate about the existence of online services that provide sexual health information and support; persuade people about their credibility; provide training, such as step-by-step instructions on how to seek information online and use online support and appraise the credibility of online information and support services; model peers successfully seeking information and using support services online; and enable use of them by ensuring they are inclusive and simple to use, including provision of providing a glossary of terms to assist with spelling for searches and communication and step-by-step instructions on how to seek information online and use online support. Ultimately, while online sexual health provision has the potential to extend access to healthcare for some, addressing the needs of underserved population outlined here is crucial to facilitate access to through the digital doorway to sexual healthcare and prevent the widening of health inequalities.

## Data Availability

Data produced in the present study are available upon reasonable request to the first author.

## Appendices

## Appendix A: A list of examples of online sexual health information and support services

| Online sexual health information/support | Website name | Website link |
| --- | --- | --- |
| **Online sexual health information** |  |  |
| Information on websites | NHS | https://www.nhs.uk/service-search/sexual-health |
|  | Brook | https://www.brook.org.uk/ |
|  | SH:24 | https://sh24.org.uk/ |
|  | Terrance Higgins Trust | https://www.tht.org.uk/ |
|  | Waverly care | https://www.waverleycare.org/ |
| **Online sexual health support** |  |  |
| Synchronous communication (live chat) with trained professional | Terrance Higgins Trust | https://www.tht.org.uk/our-services/support-services/tht-direct-helpline |
| Synchronous communication (live chat) with automated responses (chatbot) | Planned Parenthood | https://roo.plannedparenthood.org/ |
|  | PositiveEast | https://www.positiveeast.org.uk/chattopat/ |
|  | Brook | https://www.brook.org.uk/contact-us/ |
|  | NHS Blackpool Teaching Hospitals | https://www.lancashiresexualhealth.nhs.uk/sexual-health-guides/chatbot |
| Asynchronous communication (email) with a trained professional | Embrace | https://www.embracewolverhampton.nhs.uk/about-us/chat-sexual-health#:~:text=Text%20a%20sexual%20health%20nurse,Smear%20tests |
|  | Sandyford | https://www.sandyford.scot/about-us/contact-us/#:~:text=If%20you%20are%20involved%20in,scot.nhs.uk |
|  | Umbrella | https://umbrellahealth.co.uk/our-services/chathealth/ |
|  | Sexual Health Heartfordshire | https://www.sexualhealthhertfordshire.clch.nhs.uk/support/chat-sexual-health |
|  | Waverly care | https://www.waverleycare.org/home/contact-us/ |
|  | Terrance Higgins Trust | https://www.tht.org.uk/get-help/support-services/tht-direct-helpline |
|  | SH:24 | https://sh24.org.uk/contact-us |
| Asynchronous communication (SMS text) with a trained professional | Leeds Sexual Health | https://www.leedssexualhealth.com/wheretogo/chat-health |
|  | NHS Dorset HealthCare University | https://sexualhealthdorset.org/how-can-we-help/chat-sexual-health/ |

## Appendix B: An overview of seven interrelated elements of care within the behavioural system of accessing and using online sexual healthcare

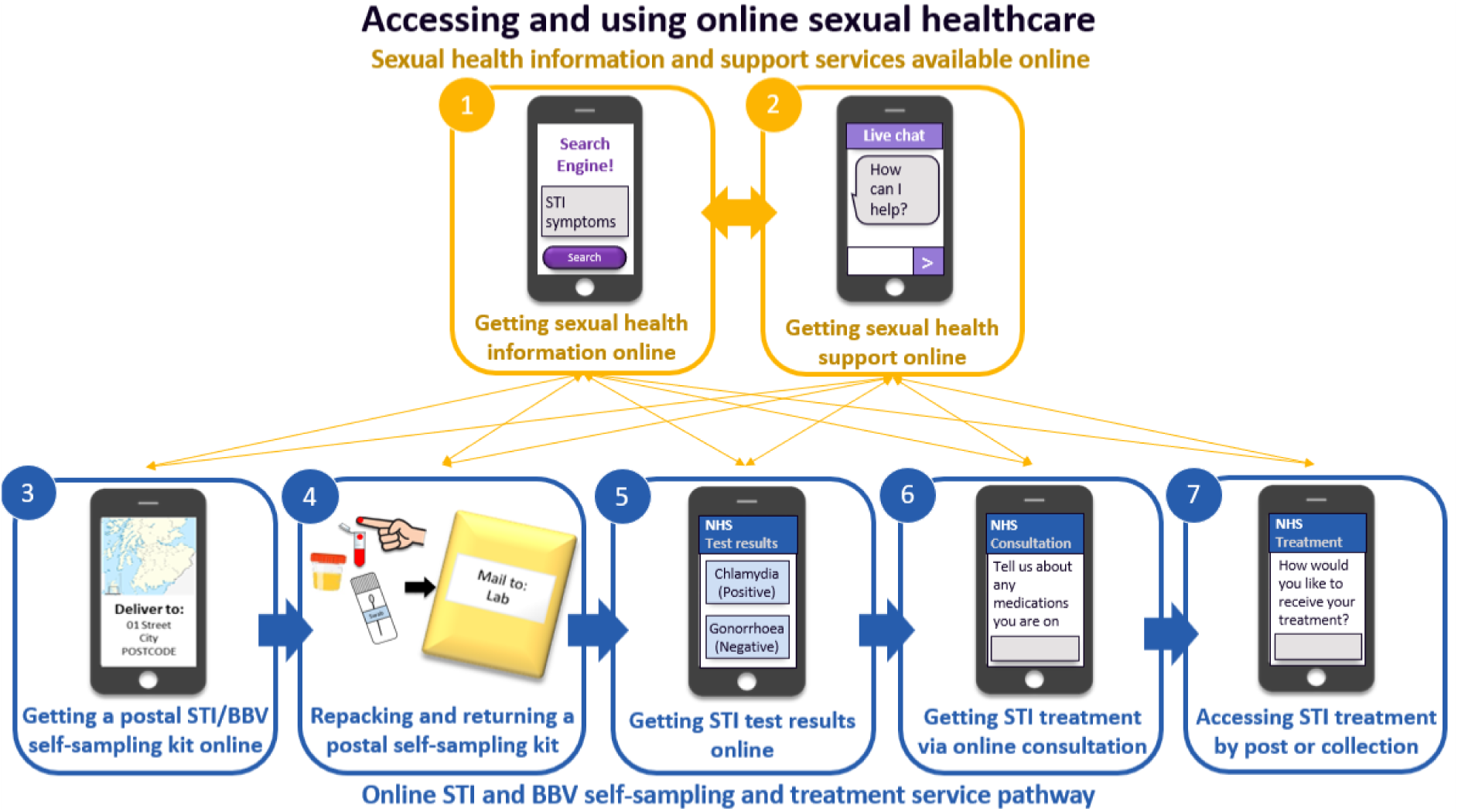
*Note.* Numbers and thick arrows represent a pathway for accessing and using online sexual healthcare, typically beginning with seeking online sexual health information and support online, which can lead to a sequential online STI and BBV self-sampling service pathway. Thick yellow arrows indicate a non-sequential order, where the elements of care can occur in any order, e.g., seeking online sexual health information can occur before, simultaneous to, or after seeking online sexual health support. Thin yellow arrows indicate that seeking online sexual health information or support can occur at any point in the STI self-sampling and treatment pathway. Thick blue arrows indicate a sequential order, where a later element of care cannot precede an earlier domain, e.g., getting STI test result online must occur after getting a postal STI/BBV self-sampling kit online.

## Appendix C: Target sampling frame developed prior to data collection

**Appendix C: Target sampling frame developed prior to data collection.**
| **Demographic characteristics and socioeconomic indicators** | **Targets (if sample n=30)** |
| --- | --- |
| **Ethnicity** | n=10 from minority ethnic groups (especially Black Caribbean, Black Other, and Mixed ethnic populations) |
| **Age** | n=15 aged under 25 years<br>n=10 aged 25 to 49 years<br>n=5 aged 50+ years |
| **Gender** | n=5-10 non-binary and trans people<br>n=20-25 cisgender |
| **Sexuality** | n=5 bisexual women<br>n=5 heterosexually identifying MSM<sup>a</sup> (HIMSM)<br>n=5-10 gay and bisexual MSM (GBMSM)<br>n=15-20 heterosexual men |
| **Disability** | n=5 with a mild learning disability |
| **Education** | n=10-15 with no higher education |
| **Employment** | n=10 out of work / in temporary work |
| **Area deprivation (IMD<sup>b</sup>)** | n=10-15 from the most deprived quintiles |
| **Geography/Area of residence** | n=5 people from a geographic area with limited access to physical sexual health services? |

<sup>a</sup>MSM = men who have sex with men
<sup>b</sup>IMD = Index of Multiple Deprivation.

## Appendix D: Socio-economic demographics and screening survey

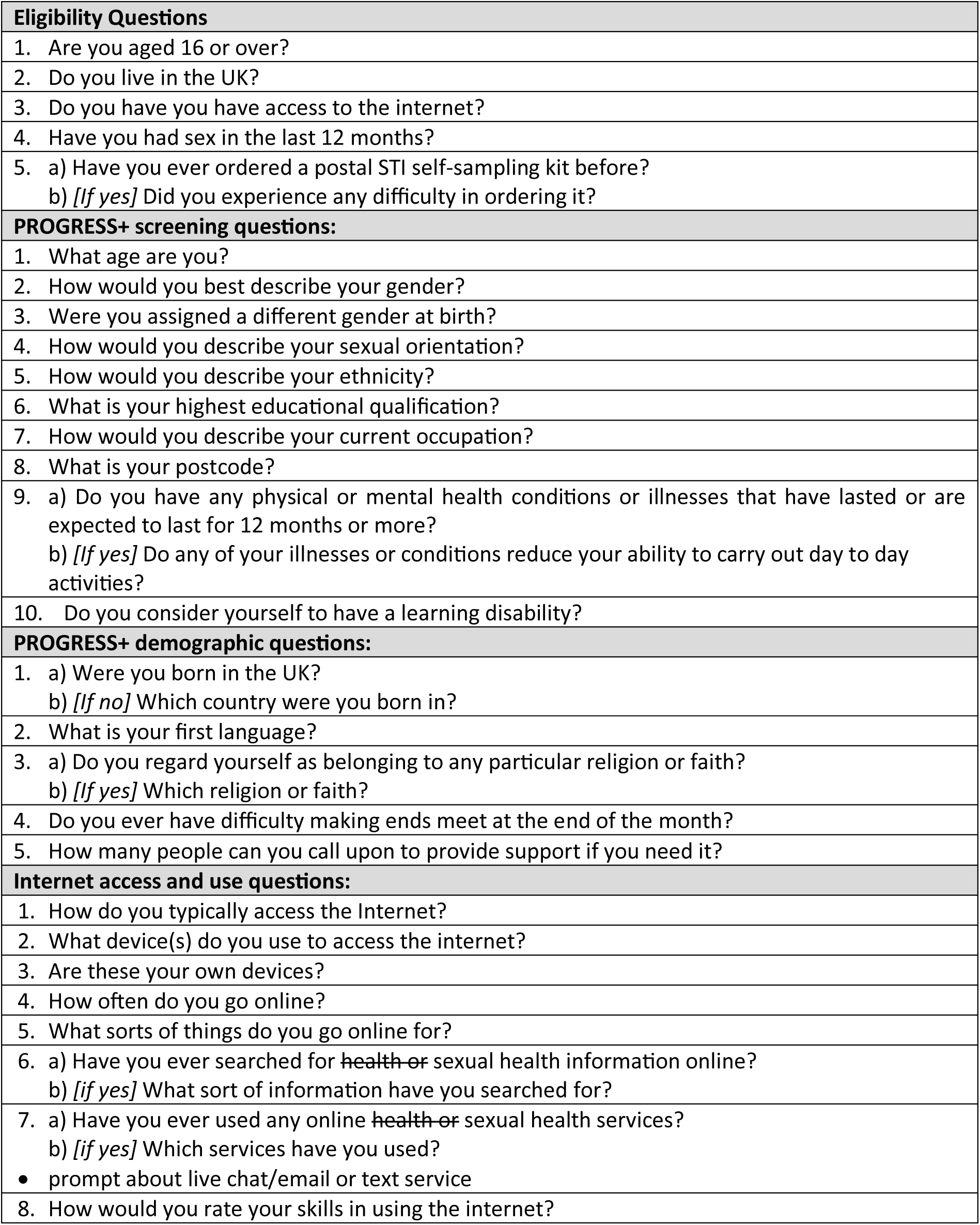

## Appendix E: Interview topic guide

Appendix E: Interview topic guide.
| Behavioural domain<br>Description of element of online sexual healthcare and corresponding slide | Specific behaviour |  | Question | Prompts |
| --- | --- | --- | --- | --- |
| **Knowledge of current online sexual healthcare (1 min)** | - |  | *[If never searched for/used SHS online before]*<br>**Can you briefly tell me about your experience of accessing sexual health services in the past?** | • How did you become aware of these health / sexual health services?<br>• What, if anything, has affected your decision not to use online sexual health services?<br>• What do you think about online sexual health services as an idea?<br>○ What do you think it may be like?<br>○ What do you think it might involve?<br>• Do you have a good idea of what STIs are? |
|  |  |  | *[If searched for/used SHS online before]*<br>**Can you briefly tell me, from your recent experience, what you know about online sexual health services/ health services?** | • How did you become aware of these online sexual health services?<br>• Can you tell me what has affected your decision to use online sexual health services? |
| **Online sexual healthcare *per se* (2 mins)** | **Access online sexual healthcare** |  | **Is there anything that might make/has made it difficult for you to access online sexual health services?** | • Is there anything that might make it difficult for you to get online to use online SHS?<br>• Think about how you normally access the internet |
|  | **Use online sexual healthcare** |  | **Is there anything that might/has make it difficult for you to use online sexual health services?** |  |
| **1. Getting sexual health information online (5-7 mins)**<br>*[#1 on slide 3 & Slide 4]*<br>You can find a wide range of information about sexual health online, on websites like the NHS and sexual health charity websites. These websites have information on them about sexual health services, including clinic locations and opening times. They also have information about sexual health, including how to have safe sex and STI symptoms and testing<br>*[Slide 5]*<br>You can search for this information on Internet, using devices like a mobile phone or computer.<br>*[Sense check]* | - | Previous experience | *[If never searched for sexual health or health information online before]*<br>**Tell me about your experience using the internet for any kind of information** | • What, if anything, has stopped you from searching for health/sexual health information online?<br>• Compared to getting sexual health information in person or on the phone |
|  |  |  | *[If only searched for other health information]*<br>**Tell me about any experience of searching for other health information online** | • What are your thoughts on searching for SH information online? |
|  |  |  | *[If have searched for sexual health information online before]*<br>**Tell me about your previous experience of searching for sexual health information online** | • What are your thoughts on searching for SH information online? |
|  |  | Barriers | **What might make/ has made it difficult for you to search for sexual health information online?** | • Why?<br>• What might you struggle with?<br>• Is there anything that might stop you/put you off?<br>• Is there anything you might be concerned about?<br>• How would you know if you could trust the website?<br>• What challenges may there be for you in searching for sexual health information online? |
|  |  | Facilitators | **What might make/has made it easy for you to search for** | • Why?<br>• What are the benefits for you to this? |
|  |  |  | **sexual health information online?** | • What might encourage you to do this?<br>• What would help you do this?<br>• What do you think would be good about it? Why? |
| **2. Getting sexual health information and advice online (5-7 mins)**<br>*[#2 on slide 3 & slide 6]*<br>Some sexual health clinic and charity websites offer a service that lets you talk to a sexual healthcare professional (doctor or nurse on the NHS) or trained staff (on a sexual health charity website). You can ask or talk about things like safe sex, sexually transmitted infections, and the services available to you. These are screenshots of these services from a sexual health website.<br>*[Slide 7]*<br>This includes a live chat service where you type your name and a question and are connected to someone immediately, or an email or text service here you type your name, email address or phone number and a question, and they respond to you by email or text as soon as possible (generally within a few hours to days)<br>*[Sense check]* | - |  | *[If never got sexual health support and advice online before]*<br>**What do you think about getting sexual health support and advice online?** | • What might be different for you about using a live chat and an email or text service to get sexual health support and advice online?<br>• Do you think there might be any difference between talking to a HCP or a trained member of staff |
|  | Live chat |  | *[If never used a live chat for getting sexual health support and advice online before]*<br>**What do you think about using a live chat service to get sexual health support and advice online?** | • Why? |
|  |  | Barriers | **What would make/ has made it difficult for you to use a live web chat to get sexual health support and advice online?** | • Is there anything you might be concerned about?<br>• Is there anything that would stop you?<br>• What might put you off using a live web chat for getting sexual health support and advice?<br>• Why wouldn't you do this?<br>• What challenges might there be for you?<br>• What might help you with this? |
|  |  | Facilitators | **What would make/ has made it easy for you to use a live web chat to get sexual** | • What do you think would encourage you to use a live web chat for getting sexual health support and advice?<br>• Why would you do this? |
|  |  |  | **health support and advice online?** | • Difference between talking to HCP and trained staff? |
|  | **Email/Text service** |  | *[If never used an email/text service for getting sexual health support and advice online before]*<br>**What do you think about using an email/text service to get sexual health support and advice online?** | • Why? |
|  |  | Barriers | **What would make/ has made it difficult for you to use an email/text service to get sexual health support and advice online?** | • What might stop you?<br>• What might put you off using an email/text service for getting sexual health support and advice?<br>• Why wouldn't you do this?<br>• What challenges might there be?<br>• What might help you with this? |
|  |  | Facilitators | **What would make/ has made it easy for you to use an email or text service to get sexual health support and advice online?** | • What would encourage you to use an email/text service to get sexual health support and advice?<br>• Why would you do this? |
|  | **Sexual health service** | Barriers |  | • Is there anything that would make it difficult for you to collect your STI treatment from a pharmacy?<br>• Challenges for you |
|  |  | Facilitators |  | • Is there anything that would make it easy for you to collect your STI treatment from a SHS?<br>• Benefits for you |

## Appendix F: Interview slides provided to every participant prior to the interview

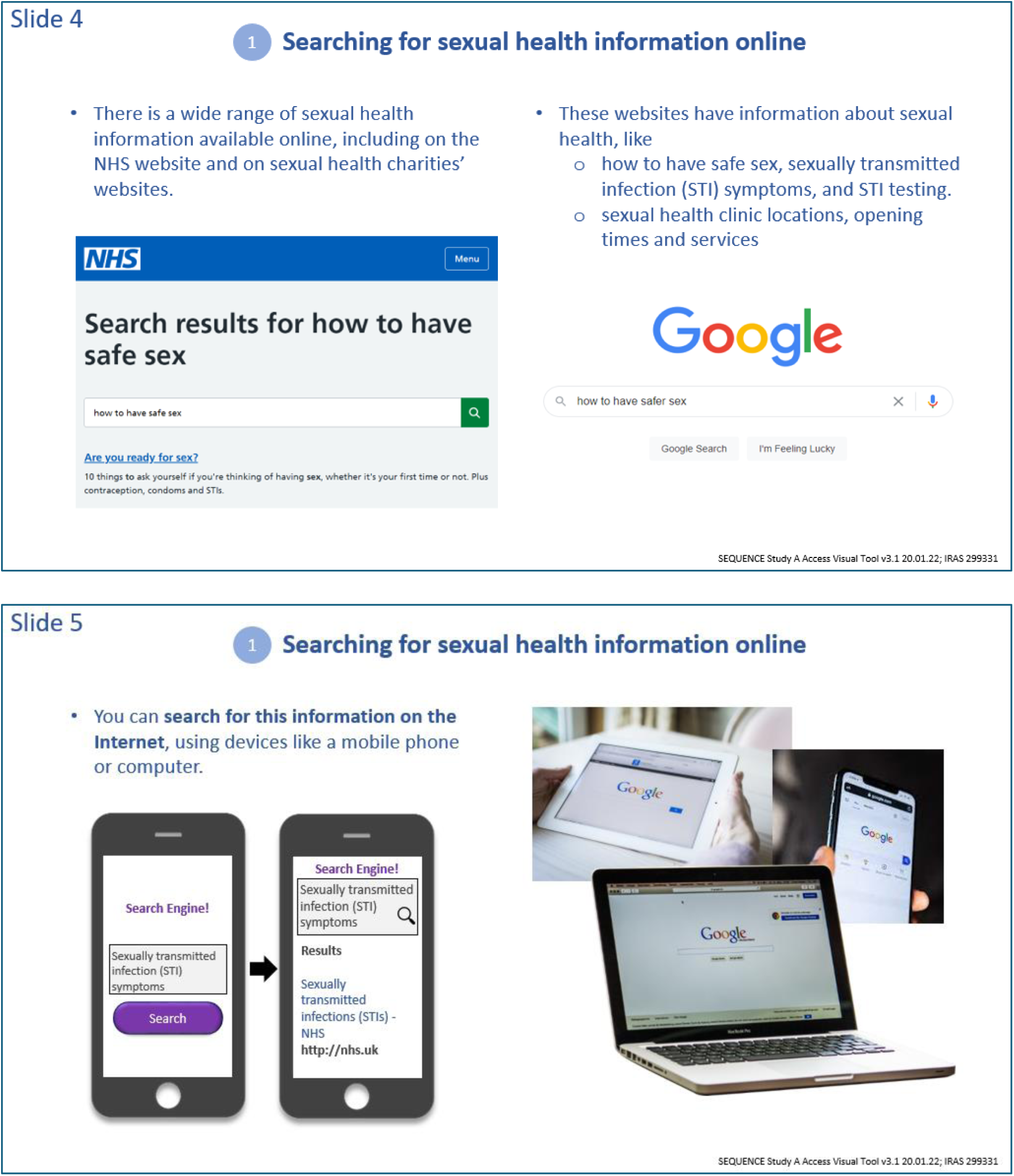
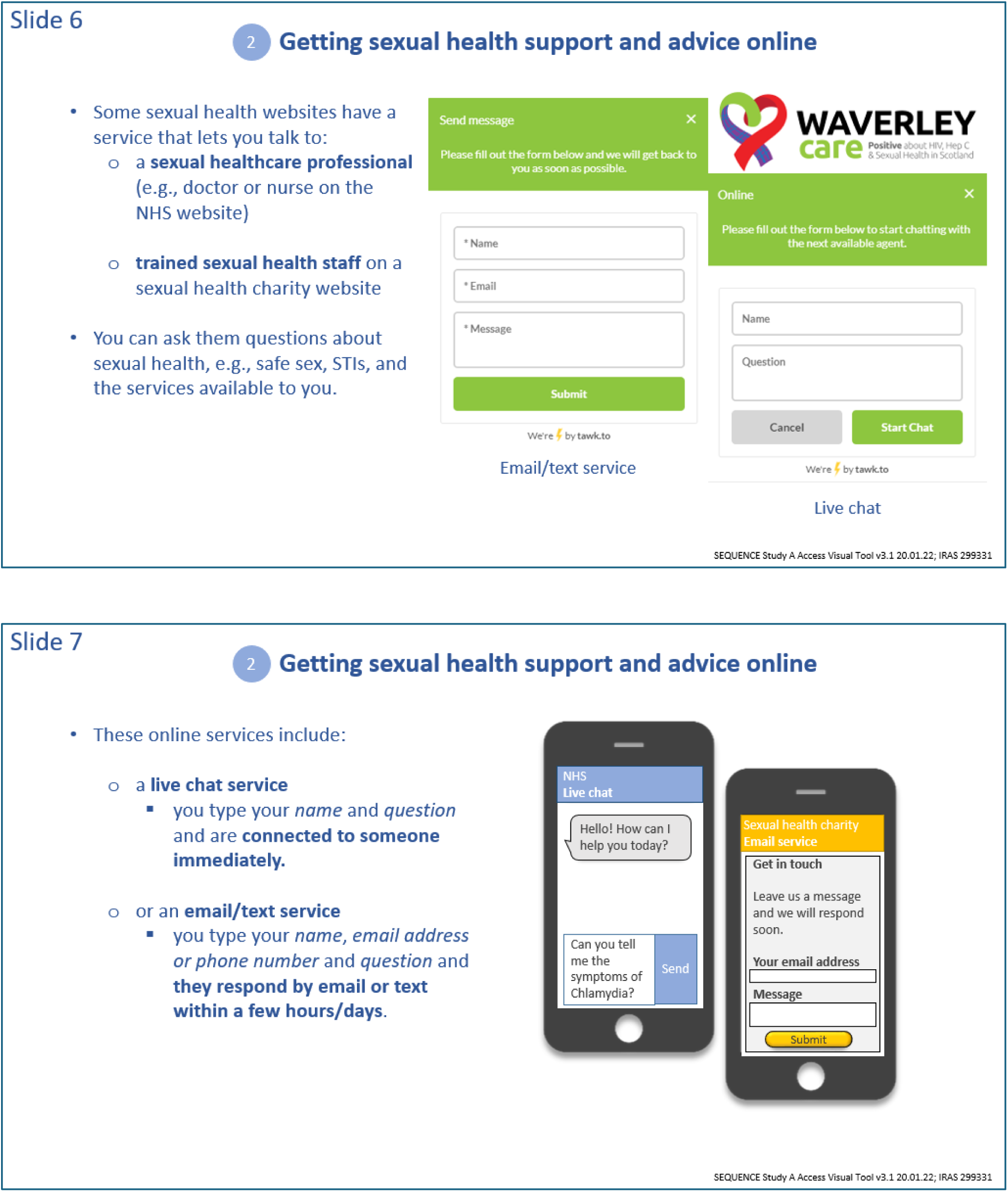

## Appendix G: Sexual health resources information provided to each participant post interview

### Sources of support

We want to make sure that you have a list of services that offer different types of support and information in case something came up in the interview that was difficult or uncomfortable for you.

### Sexual health services and organisations

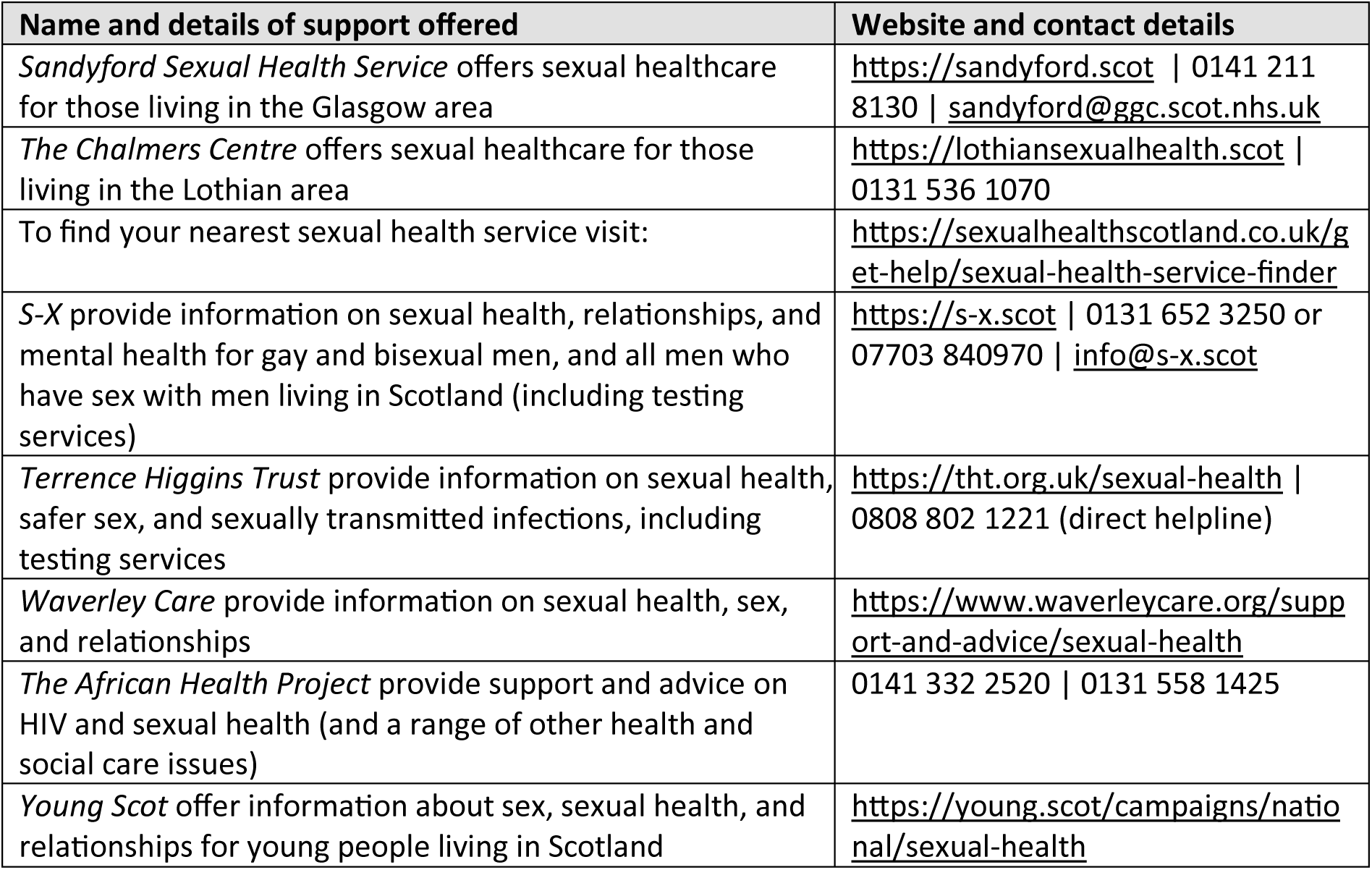

### Wider health services and organisations

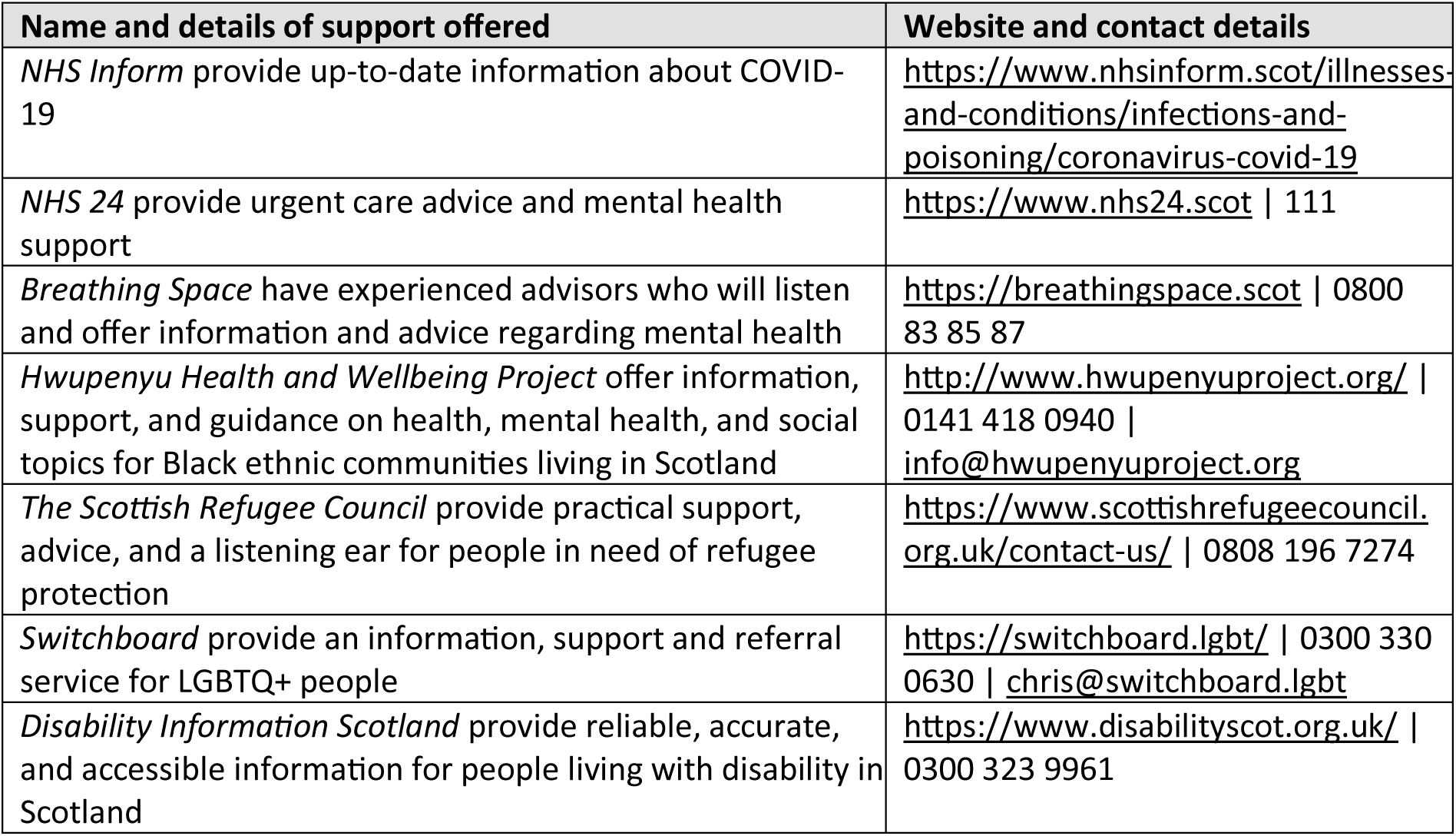
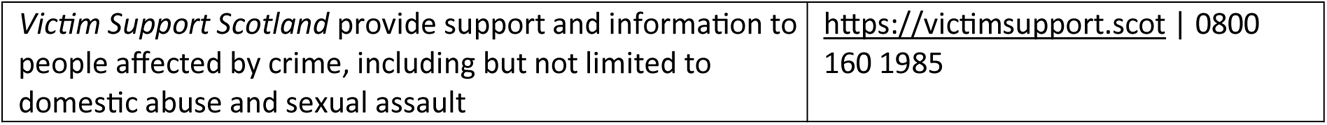

## Appendix H: Participant self-reported experience and skills using the internet

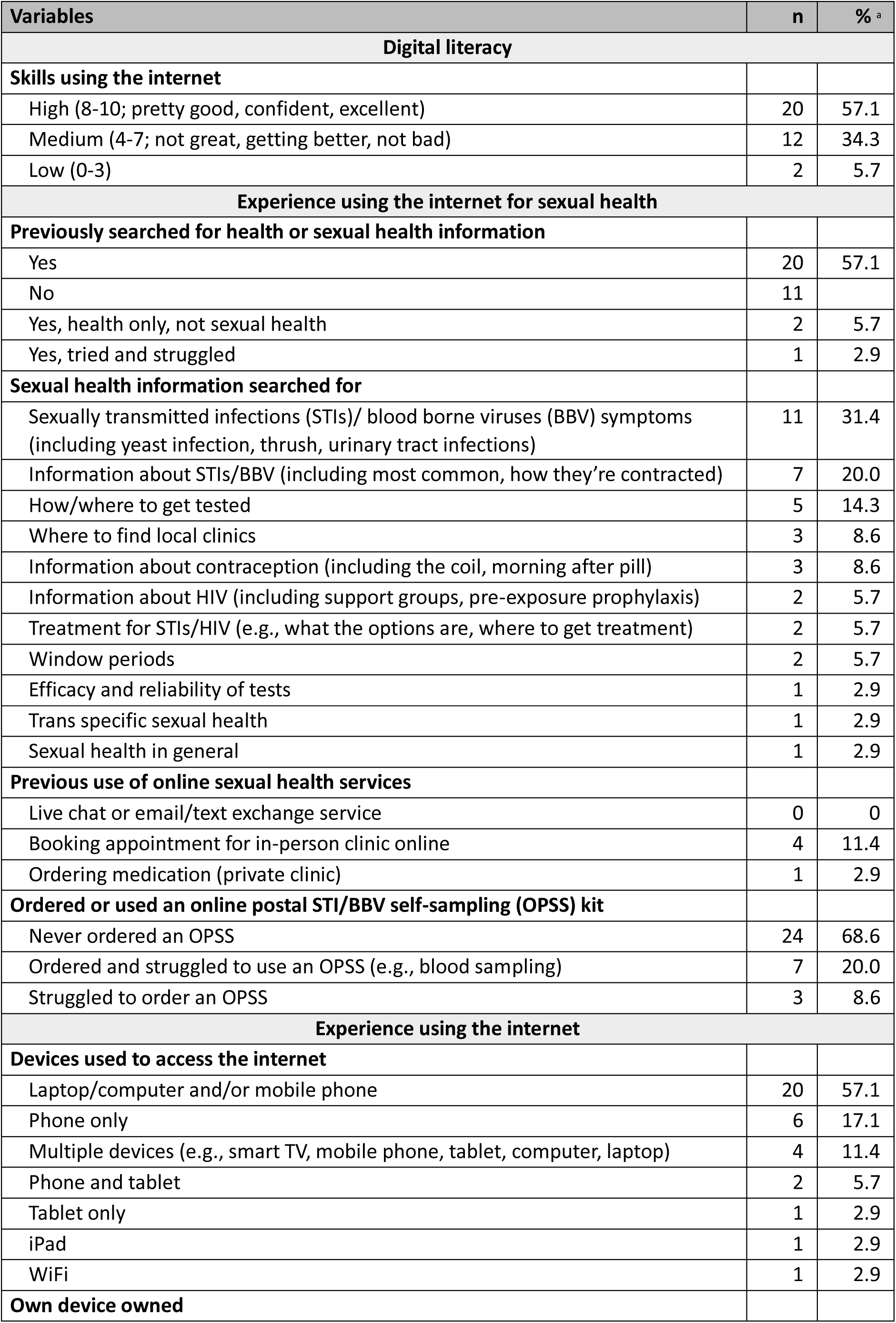
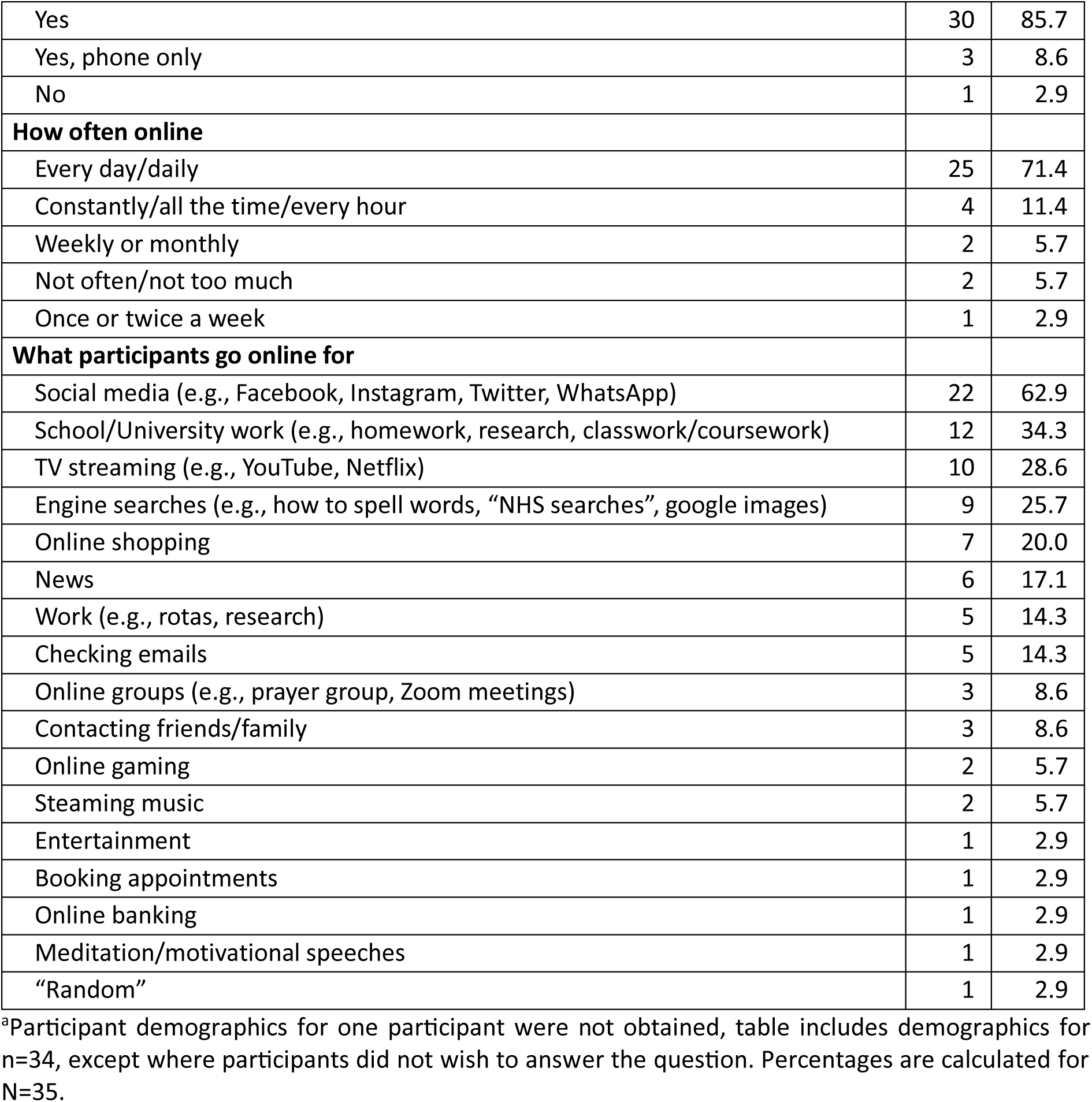

Appendix H: Participant self-reported experience and skills using the internet.
| Variables | n | %<sup>a</sup> |
| --- | --- | --- |
| **Digital literacy** |  |  |
| **Skills using the internet** |  |  |
| High (8-10; pretty good, confident, excellent) | 20 | 57.1 |
| Medium (4-7; not great, getting better, not bad) | 12 | 34.3 |
| Low (0-3) | 2 | 5.7 |
| **Experience using the internet for sexual health** |  |  |
| **Previously searched for health or sexual health information** |  |  |
| Yes | 20 | 57.1 |
| No | 11 |  |
| Yes, health only, not sexual health | 2 | 5.7 |
| Yes, tried and struggled | 1 | 2.9 |
| **Sexual health information searched for** |  |  |
| Sexually transmitted infections (STIs)/ blood borne viruses (BBV) symptoms (including yeast infection, thrush, urinary tract infections) | 11 | 31.4 |
| Information about STIs/BBV (including most common, how they're contracted) | 7 | 20.0 |
| How/where to get tested | 5 | 14.3 |
| Where to find local clinics | 3 | 8.6 |
| Information about contraception (including the coil, morning after pill) | 3 | 8.6 |
| Information about HIV (including support groups, pre-exposure prophylaxis) | 2 | 5.7 |
| Treatment for STIs/HIV (e.g., what the options are, where to get treatment) | 2 | 5.7 |
| Window periods | 2 | 5.7 |
| Efficacy and reliability of tests | 1 | 2.9 |
| Trans specific sexual health | 1 | 2.9 |
| Sexual health in general | 1 | 2.9 |
| **Previous use of online sexual health services** |  |  |
| Live chat or email/text exchange service | 0 | 0 |
| Booking appointment for in-person clinic online | 4 | 11.4 |
| Ordering medication (private clinic) | 1 | 2.9 |
| **Ordered or used an online postal STI/BBV self-sampling (OPSS) kit** |  |  |
| Never ordered an OPSS | 24 | 68.6 |
| Ordered and struggled to use an OPSS (e.g., blood sampling) | 7 | 20.0 |
| Struggled to order an OPSS | 3 | 8.6 |
| **Experience using the internet** |  |  |
| **Devices used to access the internet** |  |  |
| Laptop/computer and/or mobile phone | 20 | 57.1 |
| Phone only | 6 | 17.1 |
| Multiple devices (e.g., smart TV, mobile phone, tablet, computer, laptop) | 4 | 11.4 |
| Phone and tablet | 2 | 5.7 |
| Tablet only | 1 | 2.9 |
| iPad | 1 | 2.9 |
| WiFi | 1 | 2.9 |
| **Own device owned** |  |  |
| Yes | 30 | 85.7 |
| Yes, phone only | 3 | 8.6 |
| No | 1 | 2.9 |
| **How often online** |  |  |
| Every day/daily | 25 | 71.4 |
| Constantly/all the time/every hour | 4 | 11.4 |
| Weekly or monthly | 2 | 5.7 |
| Not often/not too much | 2 | 5.7 |
| Once or twice a week | 1 | 2.9 |
| **What participants go online for** |  |  |
| Social media (e.g., Facebook, Instagram, Twitter, WhatsApp) | 22 | 62.9 |
| School/University work (e.g., homework, research, classwork/coursework) | 12 | 34.3 |
| TV streaming (e.g., YouTube, Netflix) | 10 | 28.6 |
| Engine searches (e.g., how to spell words, "NHS searches", google images) | 9 | 25.7 |
| Online shopping | 7 | 20.0 |
| News | 6 | 17.1 |
| Work (e.g., rotas, research) | 5 | 14.3 |
| Checking emails | 5 | 14.3 |
| Online groups (e.g., prayer group, Zoom meetings) | 3 | 8.6 |
| Contacting friends/family | 3 | 8.6 |
| Online gaming | 2 | 5.7 |
| Steaming music | 2 | 5.7 |
| Entertainment | 1 | 2.9 |
| Booking appointments | 1 | 2.9 |
| Online banking | 1 | 2.9 |
| Meditation/motivational speeches | 1 | 2.9 |
| "Random" | 1 | 2.9 |

<sup>a</sup>Participant demographics for one participant were not obtained, table includes demographics for n=34, except where participants did not wish to answer the question. Percentages are calculated for N=35.

## Appendix I: Intervention Functions and their definitions, adapted from Michie et al. (45)

**Appendix I: Intervention Functions and their definitions, adapted from Michie et al. (45).**
| **Intervention Function** | **Definition of Intervention Function** |
| --- | --- |
| Education | Increasing knowledge or understanding |
| Persuasion | Using communication or induce positive or negative feelings or stimulate action |
| Training | Imparting skills |
| Modelling | Providing an example for people to aspire to or imitate |
| Environmental restructuring | Changing the physical or social context |
| Enablement | Increasing means/reducing barriers to increase capability (beyond education and training) and opportunity (beyond environmental restructuring) |

## Acknowledgements

We would like to thank the participants who took part in the study. We are also grateful to the following sexual health services and organisations for invaluable assistance with the study. We would like to thank Castle Circus Health Centre (Torbay & South Devon NHS Foundation Trust), Devon Sexual Health – Exeter & Barnstaple (Northern Devon NHS Foundation Trust), Mortimer Market Centre, Archway, Barnet and Buryfields Clinics (Central and North West London NHS Foundation Trust), Sandyford Clinic (Greater Glasgow & Clyde), and Sexual Health Sheffield (Sheffield Hospitals NHS Foundation Trust), for advertising the study to support with recruitment. We would also like to thank Hidayah LGBT and get2gether, for publicising the study to help with recruitment.

## Author contributions

**Funding acquisition:** CSE, JG, MWO, AB, PF; **Conceptualisation:** JMcL, PF, JMacD; **Project administration:** MWO; **Supervision:** PF, CSE; **Methodology:** JMacD, PF; **Resources:** JMcL, JMacD, FM; **Investigation:** JMcL; **Formal analysis:** JMcL, PF; **Visualisation:** JMcL, PF; **Writing – original draft:** JMcL, PF; **Writing – review & editing:** JMcL, PF; CSE, JMacD, JG, MWO, FM, NGM, AMD, JS, AB.

